# Hypertriglyceridemia as a Key Contributor to Abdominal Aortic Aneurysm Development and Rupture: Insights from Genetic and Experimental Models

**DOI:** 10.1101/2024.08.07.24311621

**Authors:** Yaozhong Liu, Huilun Wang, Minzhi Yu, Lei Cai, Ying Zhao, Yalun Cheng, Yongjie Deng, Yang Zhao, Haocheng Lu, Xiaokang Wu, Guizhen Zhao, Chao Xue, Hongyu Liu, Ida Surakka, Anna Schwendeman, Hong S. Lu, Alan Daugherty, Lin Chang, Jifeng Zhang, Ryan E. Temel, Y. Eugene Chen, Yanhong Guo

## Abstract

Abdominal aortic aneurysm (AAA) is a life-threatening vascular disease without effective medications. This study integrated genetic, proteomic, and metabolomic data to identify causation between increased triglyceride (TG)-rich lipoproteins and AAA risk. Three hypertriglyceridemia mouse models were employed to test the hypothesis that increased plasma TG concentrations accelerate AAA development and rupture. In the angiotensin II-infusion AAA model, most *Lpl*-deficient mice with severely high plasma TG concentrations died of aortic rupture. Consistently, *Apoa5*-deficient mice with moderately increased TG concentrations had accelerated AAA development, while human *APOC3* transgenic mice with dramatically increased TG concentrations exhibited aortic dissection and rupture. Increased TG concentrations and palmitate inhibited lysyl oxidase maturation. Locally overexpressing lysyl oxidase eliminated the impact of high TG on AAA formation in human *APOC3* transgenic mice. Administration of antisense oligonucleotide targeting *Angptl3* profoundly inhibited AAA progression in human *APOC3* transgenic mice and *Apoe*-deficient mice. These results indicate that hypertriglyceridemia is a key contributor to AAA pathogenesis, highlighting the importance of triglyceride-rich lipoprotein management in treating AAA.

**Clinical Perspective:** *What Is New?:*

- This study integrates genetic, proteomic, and metabolomic data to identify causation between increased triglyceride (TG)-rich lipoproteins and AAA risk.
- TG concentrations influence AAA formation and severity in a dose-dependent manner, potentially by inhibiting lysyl oxidase maturation and extracellular matrix assembly.
- Administration of antisense oligonucleotide targeting *Angptl3* profoundly inhibites AAA progression in human *APOC3* transgenic mice and *Apoe*-deficient mice by lowering TG concentrations.

*What Are the Clinical Implications?:*

- These findings underscore triglyceride-rich lipoprotein management as a promising therapeutic strategy for AAA treatment.
- Antisense oligonucleotide therapy targeting liver ANGPTL3 holds potential as a therapeutic approach to reduce AAA risk.

## Introduction

Abdominal aortic aneurysm (AAA) is a progressive vascular disease characterized by the localized enlargement of the abdominal aorta. The prevalence of AAA is estimated to be as high as 8% in men and 2% in women over 65 years of age.^1^ Most AAAs are asymptomatic, but the mortality rate is up to 80% if rupture occurs.^2^ As maximum aortic diameter is the most established predictor of aneurysm rupture, current guidelines recommend considering surgical repair for asymptomatic AAA patients when maximal aortic diameter is ≥ 50 mm in women and ≥ 55 mm in men. Open surgical or percutaneous endovascular aneurysm repair is still the only therapeutic option.^3^ Patients with AAA smaller than these thresholds are typically monitored through imaging surveillance. AAA-relevant risk factors, including smoking, male sex, aging, hypertension, and hyperlipidemia, are associated with enhanced aneurysm formation and growth. Currently, medications to reduce AAA risk factors, such as angiotensin II (AngII) receptor antagonists, beta-adrenoceptor blockers, statins, and anticoagulants, have not been proven effective in halting the growth of an aneurysm or preventing the rupture of asymptomatic AAA based on the findings from randomized controlled trials.^4, 5^

Our recent comprehensive genome-wide association studies (GWAS) meta-analysis, which included approximately 40,000 individuals with AAAs and over 1 million individuals without AAAs, has demonstrated that one-third of the lead variants at the 121 genome-wide significant AAA risk loci are associated with major-lipid fractions, such as total cholesterol (TC), triglycerides (TG), low-density lipoprotein cholesterol (LDL-C), or high-density lipoprotein cholesterol (HDL-C).^6^ Other genetic association studies have identified that a 1-standard deviation increase in TG concentrations is associated with a 69% higher risk of AAA.^7, 8^ Population-based studies have reported that individuals with increased serum very low-density lipoprotein-cholesterol (VLDL-C) and TG concentrations are more likely to develop larger AAA sizes and have a higher risk of rupture.^9, 10^ These epidemiologic and genetic findings suggest that increased TG-rich lipoproteins (TRL) may accelerate AAA development and rupture, and modulating TG concentrations or metabolites may become new therapeutic strategies for AAA.

TG, also known as triacylglycerols (TAG), are composed of glycerol and fatty acids and are the most abundant lipids in the body. Chylomicrons transport dietary triglycerides from the small intestines, and VLDLs transport endogenously synthesized triglycerides from the liver to peripheral tissues, such as the heart, skeletal muscle, and adipose tissues. Triglyceride metabolism is a complex process involving the breakdown and utilization of this lipid. Several proteins and enzymes play critical roles in regulating triglyceride metabolism.^11^ Among these, lipoprotein lipase (LPL), ApoC-III (apolipoprotein C-III, APOC3), ApoC-II, ApoA-V (apolipoprotein A5, APOA5), and angiopoietin-like proteins (ANGPTL3, ANGPTL4, and ANGPTL8) are key players. LPL is a critical enzyme that facilitates the hydrolysis of triglycerides carried by chylomicrons and VLDL into free fatty acids (FFA) and glycerol, allowing them to be utilized by various cells for energy or storage. Our previous study found that an intronic LPL variant, associated with increased LPL expression, was strongly related to reduced plasma TG concentrations and AAA risk.^6^ APOA5 is almost exclusively expressed in the liver and secreted in conjunction with VLDL particles. ApoC-III is a constituent of chylomicrons and VLDL particles and inhibits LPL activity, while ApoA-V and ApoC-II enhance LPL activity and promote the clearance of TG-rich lipoproteins. ANGPTL3 and ANGPTL8 are secreted proteins and inhibitors of LPL-mediated plasma TG clearance. In numerous studies involving humans and animals, loss-of-function of APOC2 or APOA5 is associated with increased plasma concentrations of TG and chylomicronemia.^12, 13^ Among the putatively causal circulating proteins associated with AAA, we identified that higher APOA5 expression is significantly associated with decreased AAA risk.^6^ Currently, gene silencing of APOC3 or ANGPTL3 significantly reduced TG and TC concentrations, making this method an effective strategy for lowering plasma lipids and preventing cardiovascular diseases.^14–17^ In the present study, we demonstrated that increased TG-rich lipoproteins accelerate AAA formation and rupture, and we provide evidence that effective management of plasma TG concentrations can alleviate AAA progression.

## Methods

The data, analytic methods, and codes will be made available to other researchers for reproducing from the corresponding author upon reasonable request. A detailed description of the methods is available in the Supplemental Materials. All genetic data used in this study were obtained from publicly available consortiums. The RNAseq original data generated during this study will be deposited in a publicly accessible database upon publication of the manuscript. All animal procedures followed the protocols approved by the Institutional Animal Care & Use Committee (IACUC) at the University of Michigan and University of Kentucky.

### Statistic analysis

Analyses of RNA sequencing and genetic data were performed using R language (v4.3.0). Statistical analyses of other data were conducted using GraphPad Prism 10. Most of the data are presented as mean ± standard error of the mean (SEM). The distribution of data was assessed using the Kolmogorov-Smirnov test (for n > 4) or the Shapiro-Wilk test (for n ≤ 4). For normally distributed data, Student’s t-tests were used to compare two groups, one-way analysis of variance (ANOVA) was used to compare multiple groups, and the Sidak method was applied for multiple comparison correction. For non-normally distributed data, Mann-Whitney U tests were used to compare two groups, Kruskal-Wallis tests were employed to compare multiple groups, and the Dunn method was used for multiple comparisons correction. A *P*-value less than 0.05 was considered statistically significant.

## Results

### Genetic, proteomic, and metabolic studies underscore high triglyceride-rich lipoproteins as a causal risk factor for AAA

Genetic variants that modulate the expression of genes encoding the candidate proteins or the abundances of metabolites represent valuable tools for guiding therapeutic strategies. Recent studies have identified cis-acting protein quantitative trait loci (cis-pQTL) that can be used as instrumental variables (IVs) for inferring causality.^18, 19^ Combining cis-pQTL and GWAS findings, we performed Mendelian randomization (MR) analyses to assess the causal effects of circulating proteins on AAA risk and identified that 41 out of 2,698 circulating proteins were significantly associated with AAA after adjusting for multiple tests (Fig 1, full MR results in Supplementary Table 1). Among them, genetically determined circulating APOC3 (OR=1.84, 95% CI [1.66-2.04]) and APOA5 (OR=0.61, 95% CI [0.56-0.67]) had the most significant effects on AAA risk. Seven of the 41 circulating proteins, including APOC3, APOA5, LPL, APOE, PLTP (phospholipid transfer protein), PCSK9 (proprotein convertase subtilisin/kexin type 9), and LPA (lipoprotein(A)), play an important role in lipid metabolism, especially in VLDL and chylomicron metabolism (Fig 1B). Using multi-instrument variable (multi-IV) MR to account for potential pleiotropy, six of the seven lipid metabolism-related proteins (APOC3, LPL, APOE, PLTP, PCSK9, and LPA) were found to have causal effects on AAA across inverse-variance weighted (IVW) MR, weighted median MR, and MR-Egger analyses (Supplementary Table 2). Our previous GWAS findings demonstrated that 42 lead variants at the AAA risk loci are associated with major lipid fractions.^6^ After prioritizing the role of five major lipoprotein components (HDL-C, LDL-C, TG, APOA1, and APOB) as risk factors for AAA, TG was the top-ranked independent risk factor for AAA with a marginal inclusion probability of 0.81 (P=0.007; Supplementary Table 3). Consistent with previous studies,^6–8^ LDL-C, HDL-C, and APOA1 still were significant factors (Supplementary Table 3). These genetic and proteomic findings indicate that TG-related metabolism may play a substantial role in AAA pathogenesis.

**Fig. 1:**
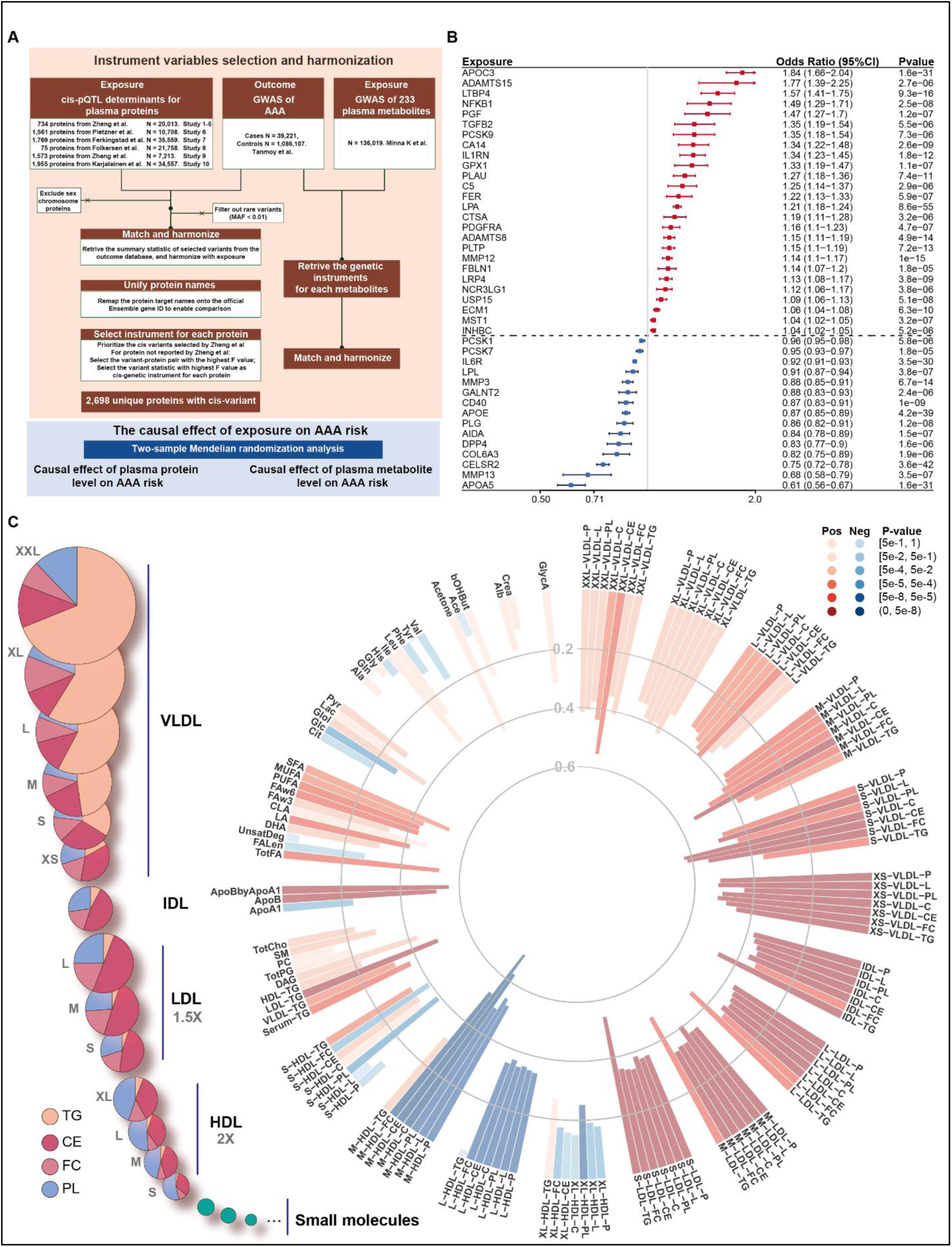
Mendelian Randomization identifies circulating proteins and metabolites causally related to AAA risk. **A**, Outline of the analyses performed. Two-sample Mendelian Randomization (MR) analyses were applied to determine the causal effects of circulating proteins, and nuclear magnetic resonance (NMR) measured circulating metabolites on the risk of abdominal aortic aneurysm (AAA) using multiple data sources. **B**, Forest plot displaying the causal effects of 41 proteins identified through MR-Wald ratio analysis after Bonferroni correction. **C**, Classifications of NMR metabolites and the relative lipid compositions of the 14 lipoprotein subclasses (left). All 14 lipoprotein subclass particles are illustrated using the same size scale, with diameters based on mean values from 5,651 participants in the Northern Finland Birth Cohort 1966 (NFBC66). Note that the size of the LDL and HDL particles in the figure is multiplied by 1.5 and 2.0, respectively. The right side features a circular histplot of the MR results for 141 metabolites. Each bar’s height represents the absolute effect size, while the color indicates the direction and P-value as determined by the MR inverse-variance weighted (IVW) method.

To further identify genetic associations between circulating metabolic traits and AAA risk, we utilized the data from a recently published GWAS study of 233 circulating metabolites to perform a two-sample MR analysis.^20^ The NMR-measured metabolites included 213 lipid traits (lipids, lipoproteins, and fatty acids) and 20 non-lipid traits (amino acids, ketone bodies, and glucose/glycolysis-related metabolites).^20^ Lipoproteins are classified by particle sizes (XXL, XL, L, M, S, and XS) and then subgrouped by the components (particle number, phospholipids, cholesterol, cholesterol ester, free cholesterol, and TG). For each metabolite, we used the IVW method as the primary MR approach and MR-Egger analysis and weighted median-based regression methods as sensitivity analyses. We demonstrated that TG-rich lipoproteins, including VLDL, IDL (intermediate density lipoprotein), and LDL, and all the components in TRL particles, were positively associated with AAA risk, while HDL and glucose were negatively associated with AAA risk, consistent with population findings (Fig 1C, full MR results in Supplementary Table 4).^9, 10, 21, 22^ Interestingly, we noticed that the TG component in HDL particles was positively associated with AAA risk compared to other components in HDL particles, such as cholesterol and phospholipids (Fig 1C). Concomitantly, we observed that total fatty acids and saturated fatty acids, the major metabolites of triglycerides, were also positively associated with AAA risk. These findings raise the possibility that lowering triglycerides and related metabolites could provide a therapeutic pathway for reducing the risk of AAA.

### Impaired LPL activity accelerates AAA development and rupture

Chronic subcutaneous infusion of AngII serves as a well-established AAA model in hypercholesterolemic mouse models, which includes using mice with deficiencies in apolipoprotein E (*Apoe*) or LDL receptor (*Ldlr*) or manipulating by adeno-associated virus-mediated overexpression of *Pcsk9* to induce hyperlipidemia.^23–25^ Our previous GWAS demonstrated that LPL variants that lead to increased plasma TG concentrations were linked to an increased risk of AAA.^6^ The MR analysis also identified circulating LPL levels to be negatively associated with AAA risk (Fig 1B). To validate whether increased TG concentrations caused by impaired LPL activity can result in AAA development and rupture, we infused AngII into male, inducible *Lpl*-deficient (i*Lpl*^-/-^) mice fed a low-cholesterol Western diet. With AngII infusion, an aortic rupture occurred in 94% (16/17, combined data from two independent studies) of i*Lpl*^-/-^ mice compared to 9.5% (2/21, combined data from two independent studies) aortic rupture rate in littermate control mice (Fig 2A-C, Supplementary Fig 1). The only survival i*Lpl*^-/-^ mouse did not have AAA and had a plasma TG concentration of 474 mg/dL and TC concentration of 233 mg/dL. In contrast, after 1 week of Western diet feeding but before AngII infusion, the majority of the i*Lpl*^-/-^ mice had severely increased TG concentrations, which were as high as 11,103 mg/dL (4,077 ± 3,218 mg/dL) compared to 66 ± 28 mg/dL in *Lpl*^f/f^ mice (Supplementary Fig 1A). Saline infusion, as a control, did not cause aortic rupture in either genotype (Supplementary Fig 1C). These findings showed that severely increased TG concentrations as a consequence of LPL deficiency resulted in aortic rupture and death in the AngII-induced AAA model.

**Fig. 2:**
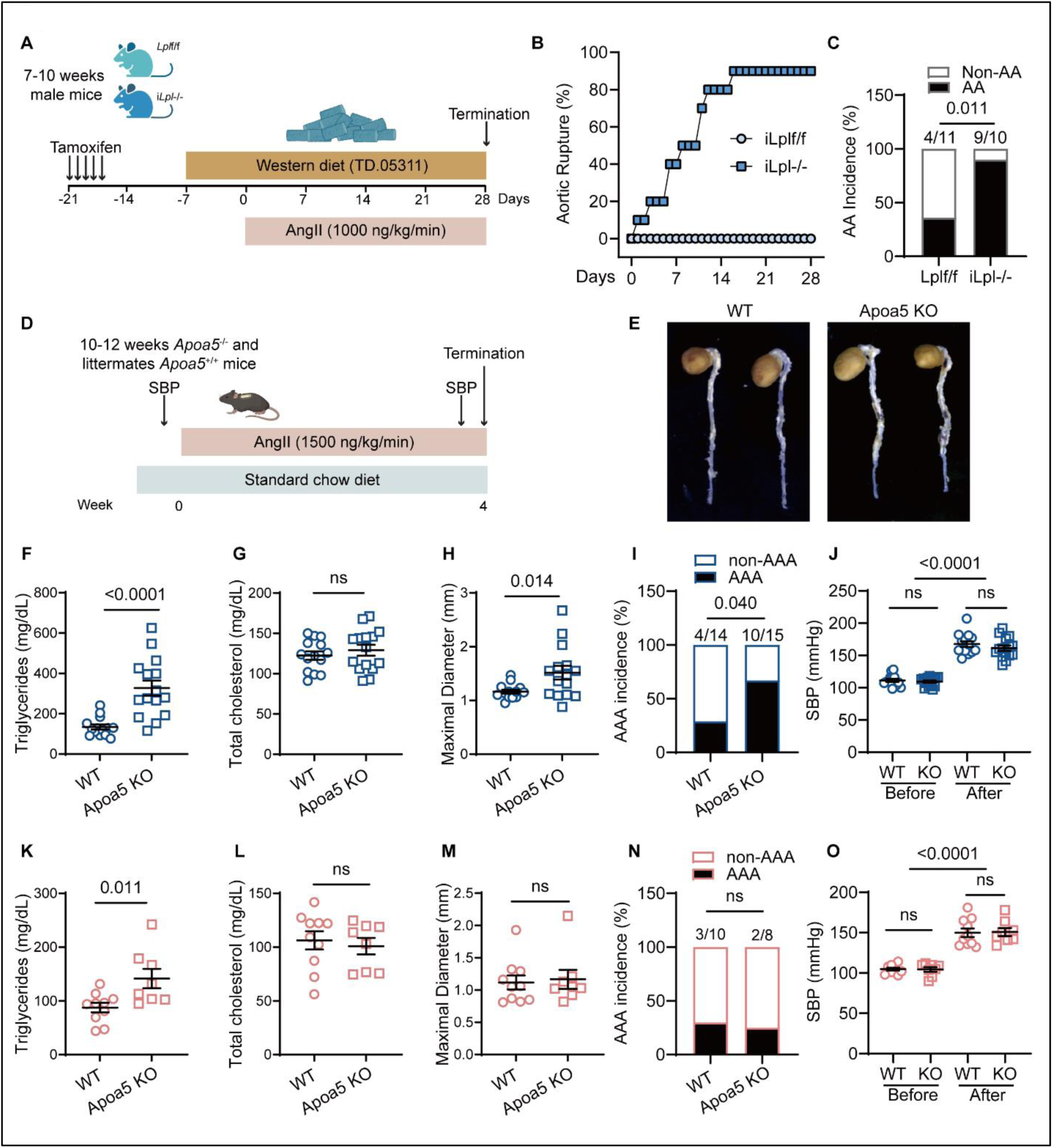
Impaired LPL activity accelerates AAA development and aortic rupture. **A**, Design of the AAA study in *Lpl*-deficient mice. Inducible global *Lpl*-deficient mice were generated by crossing *Lpl* floxed (*Lpl*^f/f^) mice with β-actin driven tamoxifen-inducible MerCreMer transgenic mice. Low cholesterol, Western diet feeding was initiated 2 weeks after the start of tamoxifen treatment. **B**, Aortic rupture incidence curve during 4 weeks of AngII infusion. **C**, Aortic aneurysm incidence in *Lpl*^f/f^ (n=11) and i*Lpl*^-/-^ (n=10) mice. **D**, Design of the AAA study in *Apoa5*-deficient mice. Mice were fed a standard rodent laboratory diet. After 4-week of AngII infusion, aortas from surviving mice were harvested. **E**, Representative aorta of male WT (n = 14) and *Apoa5*-deficient (n = 15) mice after 4 weeks of AngII infusion. Plasma samples were collected on day 28 and subjected individually to analytical chemistry to measure triglycerides (TG) and total cholesterol (TC). **F**-**J**, The TG (**F**), TC (**G**), maximal aortic diameter (**H**), AAA incidence (**I**), and systolic blood pressure (**J**) in male mice. **K-O**, TG (**K**), TC (**L**), maximal aortic diameter (**M**), AAA incidence (**N**), and systolic blood pressure (**O**) in female WT (n = 10) and *Apoa5*-deficient (n = 8) mice in AngII-induced AAA study. Data are presented as circles and Mean ± SEM (**F-H**, **J-M**, **O**). Statistical analyses were conducted as follows: Chi-Squared test for **C**, **I**, **N**; Mann-Whitney U test for **F**, **M**; Student’s t-test for **G**, **H**, **K**, **L**; Kruskal-Wallis test followed by Dunn’s post hoc analysis for **J**; One-way ANOVA followed by Sidak post hoc analysis for **O**. SBP, systolic blood pressure.

Then, we employed other mouse strains with increased, but not severely elevated, TG concentrations to validate our hypothesis. We performed all the following AAA studies in mice fed a standard rodent laboratory diet to minimize the effects of hypercholesterolemia induced by a Western diet. ApoA-V is produced in the liver and secreted into circulation, where it exerts an important role in regulating plasma TG concentrations by enhancing LPL activity to hydrolyze triglycerides. Male *Apoa5*- deficient mice showed a 2-3-fold increase in TG concentrations but no difference in TC concentrations (Fig 2D-G, Supplementary Fig 2A-B), consistent with previous findings.^12^ Using size exclusion chromatography, we found increased TG concentrations in VLDL and IDL (Supplementary Fig 2D-E) in the *Apoa5*-deficient mice compared to their littermate WT controls, while cholesterol concentrations remained similar. Plasma non-esterified fatty acids (NEFA), which are primarily released during TG hydrolysis, were also significantly elevated in the *Apoa5*-deficient mice (Supplementary Fig 2C). AAA was evaluated after a 4-week AngII infusion. The hypertriglyceridemic *Apoa5*-deficient mice had increased AAA incidence and larger maximal abdominal aorta diameter (Fig 2E, H, I). Slightly increased TG concentrations in *Apoa5*-deficient mice promoted AAA formation, but did not trigger aneurysm rupture and animal death. There was no difference in blood pressure before and after AngII infusion between *Apoa5*-deficient mice and their littermate controls (Fig 2J). This phenotype was not observed in *Apoa5*-deficient female mice (Fig 2K-O), which had significantly lower TG concentrations compared to male *Apoa5*-deficient mice (142 ± 51 mg/dL v.s. 327 ± 147 mg/dL, *p* = 0.003, Fig 2F and K). In addition, female mice demonstrate markedly lower incidences of AAA than males due to sex hormones or sex chromosomes.^26^ These findings imply the causative roles of elevated TG concentrations in AAA pathogenesis.

### Moderately to severely increased triglyceride levels accelerate AAA development and dissection in human *APOC3* transgenic mice

Increased plasma ApoC-III concentrations are linked to enhanced production and slowed clearance of triglyceride-rich lipoproteins, causing hypertriglyceridemia. Human *APOC3* transgenic (h*APOC3* Tg) mice and age/sex-matched wild-type littermates were fed a standard rodent laboratory diet (Fig 3A). Compared with controls, male h*APOC3* Tg mice had about 8-fold increase in plasma TG concentrations (Fig 3B) and about 1-fold elevations in plasma TC concentrations (Fig 3C) and non-esterified fatty acids (NEFA, Fig 3D). We observed a low incidence of AAA in wild-type littermates (2 out of 17), similar to the literature.^26^ However, there was a dramatically increased AAA incidence and a larger maximal diameter of the suprarenal aorta in h*APOC3* Tg mice (11 out of 13 surviving animals), as well as a higher dissection rate (8 out of 13 surviving animals) and another 2 mice dying from AAA rupture (Fig 3E-H, Supplementary Fig 11). There was no difference in systolic blood pressure (SBP) between h*APOC3* Tg mice and controls (Fig 3I). A similar phenotype was observed in female mice (Fig 3J-O), which had 5-fold increase in TG concentrations compared to control mice, indicating that dramatically increased TG concentrations accelerated AAA development, surpassing the protective effects of female hormones.

**Fig. 3:**
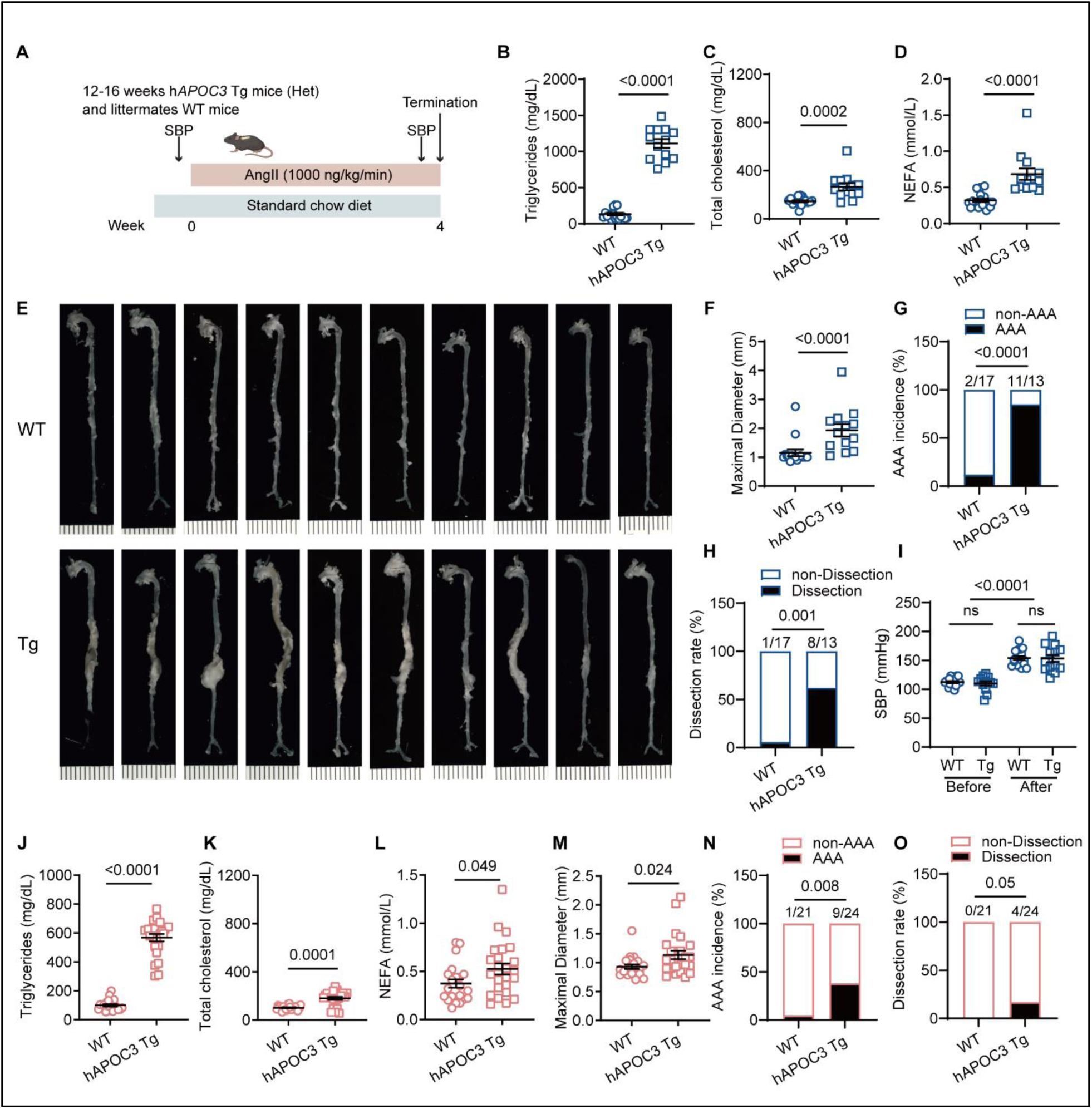
Dramatically increased TG concentrations accelerate AAA development and dissection in h*APOC3* Tg mice. **A**, Design of the AAA study in human *APOC3* transgenic (h*APOC3* Tg) mice. Twelve- to 16-week-old h*APOC3* Tg mice (n of male = 15; n of female = 24) or WT littermates (n of male = 17, n of female =21) were infused with AngII (1,000 ng/kg/min) for 4 weeks to induce AAA. Systolic blood pressures were measured before and after AngII infusion. The mice were fed a standard rodent laboratory diet. After 4 weeks, aortas from surviving mice were harvested. Plasma samples were collected on day 28 and subjected individually to analytical chemistry. **B**-**D**, plasma triglycerides (TG) (**B**), total cholesterol (TC) (**C**), and non-esterified fatty acids (**D**) in male mice. Representative aortas of male WT and h*APOC3* Tg mice after 4 weeks of AngII infusion (**E**). Maximal aortic diameter (**F**), AAA incidence (**G**), dissection rate (**H**), and systolic blood pressure (**I**) in male mice. **J**-**Q**, The TG (**J**), TC (**K**), non-esterified fatty acids (**L**), maximal aortic diameter (**M**), AAA incidence (**N**), and dissection rate (**O**) in female WT and h*APOC3* Tg mice after 4 weeks of AngII infusion. Data are presented as circles and Mean ± SEM (**B-D**, **F**, **I-M**). Statistical analyses were conducted as follows: Mann-Whitney U test for **B**, **D**, **F**, **J**, **K**; Student’s t-test for **C**, **L**, **M**; Chi-Squared test for **G**, **H**, **N**, **O**; One-way ANOVA followed by Sidak post hoc analysis for **I**. Scale bars: 1 mm in **E**. SBP, systolic blood pressure; NEFA, non-esterified fatty acids.

The effects of increased TG concentrations on AAA development were also evaluated in a PPE (porcine pancreatic elastase)-induced AAA model, in which AAA progression is independent of hypercholesterolemia.^27^ Based upon a 50% increase in the maximal abdominal aorta diameter as AAA on day 14 post-operative, the incidence of AAA was 100% for both male and female h*APOC3* Tg and littermate control mice (Supplementary Fig 3 and Supplementary Fig 4). Male h*APOC3* Tg mice showed significantly larger maximal aortic diameters than littermate controls, demonstrating that increased TG concentrations accelerated AAA growth (Supplementary Fig 3A-E). Even though female h*APOC3* Tg mice had lower TG concentrations than males (635 ± 36 mg/dL v.s. 946 ± 258 mg/dL, *p* = 0.0005), the increased TG concentrations in females were sufficient to cause larger aortic diameters (Supplementary Fig 4A-D).

These findings from the three hyperglyceridemia mouse models indicate that increased TG concentrations accelerate AAA rupture and aneurysm growth.

### Palmitate and increased triglyceride concentrations inhibit lysyl oxidase maturation in human aortic smooth muscle cells and aortas

The pathophysiology of AAA is intricately linked to the disruption of smooth muscle cell (SMC) homeostasis within the aortic wall, including phenotypic switching, cell death, extracellular matrix (ECM) remodeling, and inflammatory responses.^28^ Serum from *Apoa5*-deficient or h*APOC3* Tg mice, as well as VLDL and HDL particles, did not affect primary human aortic smooth muscle cell (HASMC) viability (Supplementary Fig 5)). Overexpression of APOC3 in h*APOC3* transgenic mice caused elevated plasma TG concentrations accompanied by increased plasma NEFA concentrations (Fig 3D and Supplementary Fig 3C). The high availability of fatty acids in the liver further increases triglyceride synthesis, resulting in VLDL assembly and secretion. Using MR, we confirmed the causal association between genetically determining plasma TG level and circulating total fatty acid concentrations (Fig 4A). Palmitic acid (PA, 16:0), stearic acid (18:0), palmitoleic acid (16:1n-7), and oleic acid (18:1n-9) are major saturated and monounsaturated fatty acids that affect cellular signaling and metabolic pathways.^29^ PA is the most common saturated fatty acid in the human body, typically accounting for 20–30% of the total fatty acids.^30^ MR analysis found that increased TG causally increased PA level, but not the other three fatty acids (Fig 4A), suggesting PA serves as an important metabolite in mediating the AAA-promoting effects of high TG or high TG-related mutations. Using untargeted metabolomics, we found elevated plasma ethyl palmitate levels (a lipid-soluble form of PA) in h*APOC3* Tg mice (Fig 4B). Bulk RNA sequencing (RNA-seq) analysis was performed to compare PA-incubated HASMC with BSA-treated (vehicle control) cells. Compared to the BSA-incubated group, PA- incubated HASMC showed an apparent reduction in ECM assembly pathways, including collagen chain trimerization, elastic fiber formation and maturation (Fig 4C-E), and increased expression of the inflammation-related gene (Supplementary Fig 6A-B, Supplementary Table 5).

**Fig. 4:**
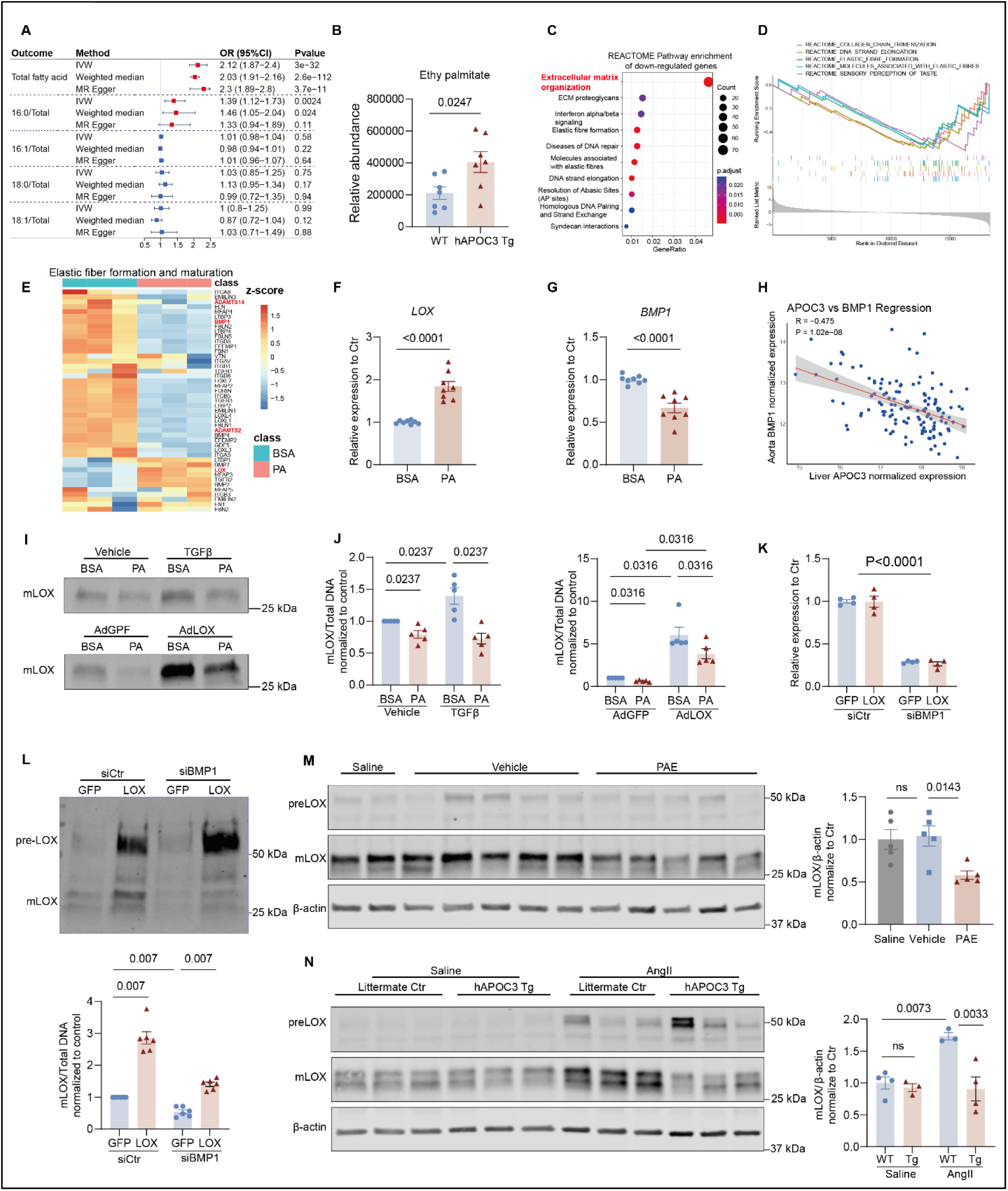
Palmitic acid inhibits LOX maturation in HASMCs and aortas. A, Two-sample mendelian randomization to detect the causal effects of TG on circulating total fatty acid, palmitic acid (PA, 16:0), stearic acid (18:0), palmitoleic acid (16:1n-7), and oleic acid (18:1n-9) levels. **B**, Untargeted metabolomics was applied to measure the ethyl palmitate levels in plasma from 14-18-week-old male h*APOC3* Tg (heterozygous) mice and littermate controls. **C-G**, Human aortic smooth muscle cells (HASMCs) were starved in OptiMEM-reduced serum medium for 24 hours, followed by incubation with palmitic acid (250 µM; PA) or vehicle (BSA) for another 24 hours. Total RNA was extracted for RNA sequencing or qRT-PCR. **C,** Reactome pathway enrichment analysis of down-regulated genes in the PA group. **D**, Top 5 downregulated Reactome pathways identified from Gene Set Enrichment Analysis (GSEA). **E**, Heatmap of gene expression levels involved in elastic fiber formation and maturation. qRT-PCR analysis of *LOX* (**F**) and *BMP1* (**G**) (data from 4 independent experiments). **H**, Correlation between liver *APOC3* expression level and aortic *BMP1* expression level among 131 donors in the GTEx project. **I-J**, HASMCs were starved in OptiMEM for 24 hours, then incubated with PA (250 µM) or vehicle along with TGF-β (10 ng/ml) or vehicle for another 24 hours; Or, HASMCs were transfected with adenovirus GFP or LOX (30 MOI) for 6 hours in a growth medium, then starved in OptiMEM for another 24 hours. Cells were incubated with PA (250 µM) or vehicle for an additional 24 hours. Conditioned medium was collected for Western blot analysis. Cell total DNA was used for normalization. **I**, Representative Western blot image of mature LOX in conditioned medium. **J**, Quantification analysis of mature LOX protein abundance. **K-L,** HASMC were transfected with siBMP1 at 1 nM or siCtr with RNAimax for 6 hours in OptiMEM-reduced serum medium, then transfected with adenovirus GFP or LOX (30 MOI) for another 23 hours in OptiMEM-reduced serum medium. The condition mediumed was used for Western blot analysis and whole cells were used for RNA extraction or DNA extraction. **K**, qRT-PCR analysis of *BMP1*. **L**, Representative LOX expression in conditioned medium (upper) and quantification analysis (bottom). **M**, Eight-week-old male C57BL/6J mice were given saline, vehicle, or ethyl palmitate (600 mg/kg) for 5 consecutive days via intraperitoneal injection. On day 6, mice were euthanized, suprarenal abdominal aortas were isolated, and total protein was extracted and analyzed by Western blot to detect LOX abundance in suprarenal abdominal aortas, with corresponding quantifications for mature LOX (n = 5/group). **N**, Twelve- to 16-week-old h*APOC3* Tg mice and littermate control mice were infused with saline or AngII (1,000 ng/kg/min) for 7 days. On day 8, mice were euthanized, suprarenal abdominal aortas were isolated, and total protein was extracted and analyzed by Western blot to detect LOX expression in suprarenal abdominal aortas, with corresponding quantifications for mature LOX (n = 3 or 4/genotyping/treatment). Data are presented as dots and Mean ± SEM (**B**, **F-G**, **J-N**). Statistical analyses were conducted as follows: One-way ANOVA followed by Sidak post hoc analysis for **K, M, N**. Student’s t-test for **B, F**, **G**; Mann-Whitney U test followed by Bonferroni correction for **J, L.**

TGF-β and lysyl oxidase (LOX) are crucial in regulating the cross-linking of collagen and elastin in the ECM. LOX was unexpectedly upregulated compared to the most downregulated genes in the elastic fiber formation and maturation pathways (Fig 4E). In primary HASMC, PA incubation induced the transcription of LOX, showing increased mRNA abundance by qRT-PCR (Fig 4F). LOX is initially synthesized as a preproprotein, which undergoes several post-translational modifications for its maturation and functional activation. Impairments in LOX activity, whether due to genetic mutations or LOX inhibitors, can lead to AAA formation and rupture.^31–33^ Our RNA-seq data indicated that PA incubation downregulated the expression of BMP-1 (Bone morphogenetic protein 1), ADAMTS2 (a disintegrin and metalloproteinase with thrombospondin motif 2), and ADAMTS14, which are critical enzymes controlling LOX activation by the proteolytic removal of the propeptide region.^34^ We further confirmed that PA incubation downregulated BMP-1 and ADAMTS2 transcriptions in HASMC (Fig 4G, Supplementary Fig 7A). Immunoblot analyses demonstrated that incubation with TGF-β increased the abundance of the mature form of LOX both in the conditioned medium and cell lysates of HASMC. However, the presence of PA blocked its effect, resulting in a less mature LOX (Fig 4I, J, Supplementary Fig 7H). LOX overexpression increased the mature form of LOX in HASMC, which was suppressed by incubation with PA (Fig 4I,J, Supplementary Fig 7I), indicating that PA interferes with the maturation process of LOX in HASMC.

To investigate how PA influences LOX maturation, using siRNA-mediated gene silence of *BMP1*, *ADAMTS2*, or *ADAMTS14* in HASMC, we found that *BMP1* knockdown led to dramatically decreased LOX maturation in the conditioned medium (Supplementary Fig 7E-F), suggesting that BMP-1 is the primary enzyme regulating LOX maturation in HASMC. Additionally, *BMP1* knockdown increased LOX transcription (Supplementary Fig 7G), likely as a compensatory effect; however, it was insufficient to compensate for the loss of mature LOX. Next, we investigated whether LOX overexpression can restore mature LOX levels within the PA-BMP-1 axis. In the presence of PA or knockdown of *BMP1*, LOX maturation was largely impaired (Figure 4I and L), showing dramatically reduced mature bands. Overexpression of LOX restored mature LOX levels (Figure 4I and L), suggesting that LOX overexpression may prevent the pro-AAA effects of high TG and PA levels. Our previous study validated the utility of tissue-crosstalk analysis for investigating organ-organ interactions.^35^ Among the >1,800 secreted proteins in the liver, APOC3 ranked 52^nd^ in terms of its impact on aortic gene expression. Among the 131 donors from the GTEx project, liver *APOC3* RNA expression showed a strong negative association with *BMP1* RNA expression in the aorta (Fig 4H), whereas the associations with *ADAMTS2* and *ADAMTS14* were much weaker (Supplementary Fig 7B-C). In contrast, liver *APOC3* expression was positively associated with aortic *LOX* expression (Supplementary Fig 7D). The tissue-crosstalk analysis suggests that liver-secreted APOC3 regulates aortic gene expression, including BMP1 and LOX, essential for vascular integrity, potentially influencing aortic extracellular matrix remodeling and contributing to AAA pathogenesis.

To investigate the effects of increased circulating palmitate concentrations on the maturation of LOX in aortas, we administered ethyl palmitate, which can be hydrolyzed to free palmitate in rodents.^36^ The mature LOX abundance in suprarenal abdominal aortas was decreased significantly after administration of ethyl palmitate compared to the vehicle or saline control aortas (Fig 4M). Then, we examined whether increased TG concentrations affected the maturation of LOX in the aortas of h*APOC3* Tg and littermate control mice. There was no difference in the abundance of mature LOX in suprarenal abdominal aortas between the two groups without AngII infusion (Fig 4N). We observed a dramatic upregulation of mature LOX upon AngII infusion in control mice (Fig 4N), suggesting a protective and compensatory mechanism to enhance elastic fiber assembly to protect against the effects of AngII. However, this response was absent in h*APOC3* Tg mice (Fig 4N), indicating that increased TG and/or palmitate inhibited LOX maturation in the aortas, which may contribute to AAA development and rupture.

### LOX overexpression effectively prevents AAA formation and dissection in h*APOC3* Tg Mice

These findings suggest that TG and palmitate promote AAA development by disrupting essential LOX maturation. To validate this mechanism *in vivo*, we utilized an adenoviral vector to overexpress LOX in the suprarenal abdominal aorta of both WT and h*APOC3* Tg mice, followed by an 18-day AngII infusion (Fig 5A). First, we confirmed that local adenovirus application successfully increased the expression of LOX or GFP (as control) in the target region (Supplementary Fig 8). LOX overexpression (LOX OE) did not affect the rupture rate (Supplementary Fig 11) or induce AAA formation in WT mice (Fig 5B-E). Strikingly, none of the h*APOC3* Tg mice developed AAA when LOX was overexpressed locally alongside AngII induction, compared to a 67% AAA incidence and a 44% dissection rate in the GFP-overexpressed control group (Fig 5B-D). LOX OE significantly reduced the maximal suprarenal abdominal aorta diameter (Fig 5E), without affecting SBP (Fig 5F), body weight (Fig 5G), TG (Fig 5H), TC (Fig 5I), and NEFA levels (Fig 5J). Histological analysis revealed that h*APOC3* Tg mice exhibited substantial collagen disorganization and elastic fiber degradation compared to WT mice, indicating impaired LOX function (Fig 5K). Notably, LOX OE restored these structural features and reinforced vascular wall integrity, even in the presence of elevated TG levels (Fig 5K). Taken together, these results established that impaired LOX activation is a key mechanism by which elevated TG levels contribute to AAA development.

**Fig. 5:**
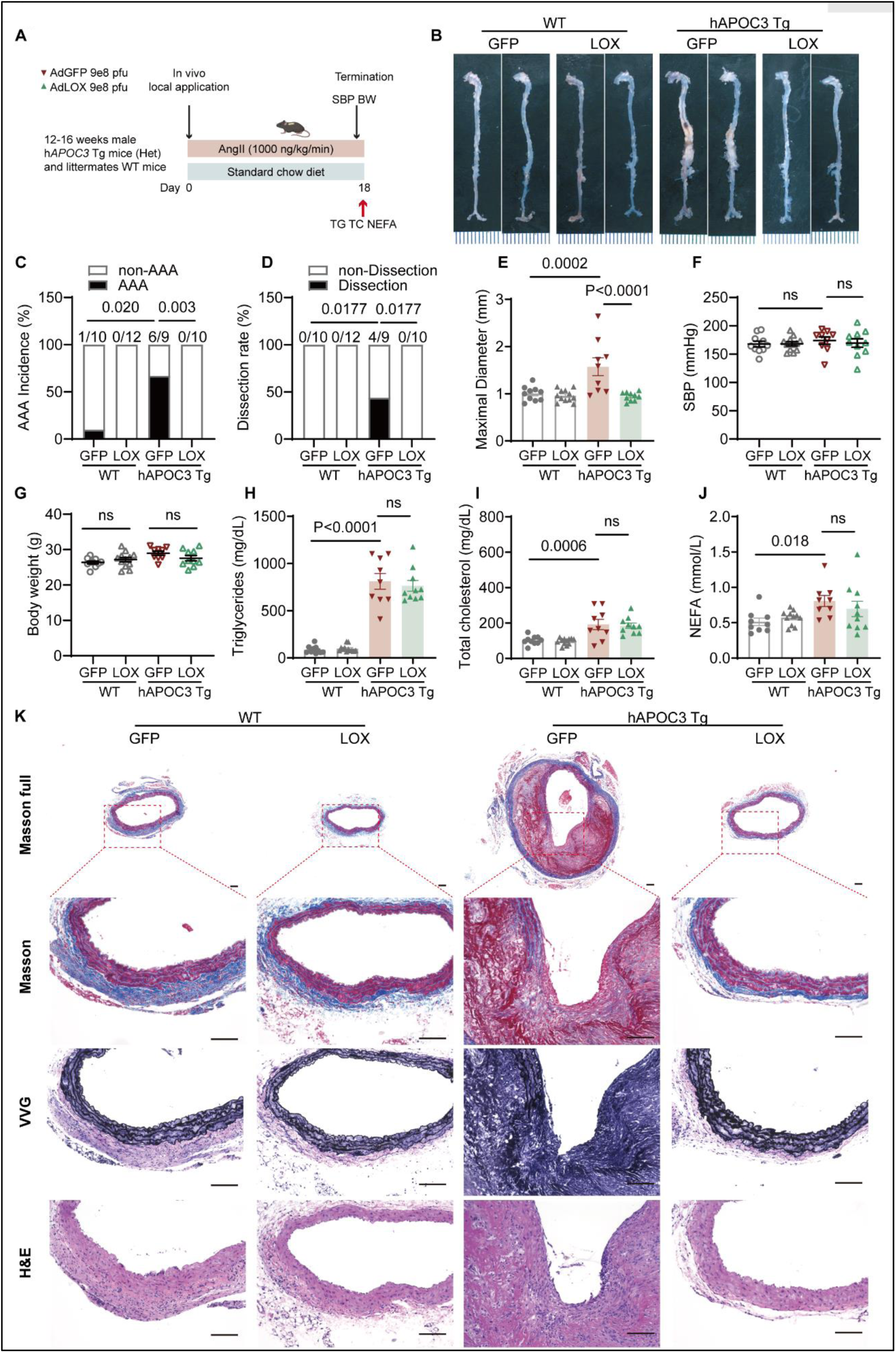
LOX Overexpression Inhibits AAA formation and dissection in *hAPOC3* Tg Mice. **A**, Design of the LOX overexpression (OE) study in human *APOC3* Tg mice. Twelve- to 16-week-old male h*APOC3* Tg mice and their littermates WT controls were transfected with 9×10^8^ pfu adnovirus GFP or LOX to the suprarenal abdominal aorta then infused with AngII (1,000 ng/kg/min) for 18 days to induce AAA. **B**, Representative aortic trees from the 4 groups. **E-F**, Quantifications of AAA incidence (**E**), dissection rate (**F**), and maximal aorta diameter (**G**) among the 4 groups. Systolic blood pressure (**H**), body weight (**I**), plasma triglycerides (TG) (**J**), total cholesterol (**K**), and non-esterified fatty acids (**L**). **M**, Representative Masson’s trichrome, Verhoeff-Van Gieson (VVG), and H&E staining of suprarenal abdominal aorta sections. Data are presented as dots and Mean ± SEM (**E-J**). Statistical analyses were conducted as follows: Chi-Squared test for **C, D**; One-way ANOVA followed by Sidak post hoc analysis for **E-G, I-J;** Kruskal-Wallis test followed by Dunn’s post hoc analysis for **H**. Scale bars: 1 mm in **B**, 100 µm in **K**.

### *Angptl3* ASO inhibits AAA formation and dissection in *APOC3* Tg mice

Based on the above findings, we asked whether lowering TG levels could attenuate AAA formation and dissection. We administrated fenofibrate and niacin, commonly used for treating hypertriglyceridemia, but only observed a 14-17 % reduction of plasma TG concentrations in h*APOC3* Tg mice (Supplementary Fig 9). N-acetylgalactosamine (GalNAc)-conjugated antisense nucleotides (ASO) targeting of hepatocyte ANGPTL3 results in significantly decreased plasma TG concentrations in animals and humans.^15, 16, 37^ In h*APOC3* Tg mice, administration of *Angptl3* ASO dramatically suppressed hepatic *Angptl3* mRNA abundance by 64% and the plasma ANGPTL3 concentrations by 81% (Fig 6A-C). Consequently, administration of the *Angptl3* ASO significantly reduced concentrations of NEFA, triglycerides (from 1,119 ± 98 mg/dL to 586 ± 114 mg/dL) and total cholesterol (from 283 ± 103 mg/dL to 117 ± 58 mg/dL) (Fig 6D-F). Consistent with the above findings, increased TG concentrations in h*APOC3* Tg mice accelerated AAA development during AngII infusion (Fig 6G-L). After administration of *Angptl3* ASO to h*APOC3* Tg mice, we recorded significant reductions in AAA incidence, maximal aortic diameter, elastin degradation, and dissection, comparable to the control mice (Fig 6G-L). qRT-PCR and ELISA analysis showed no significant changes in the levels of mouse *Apoc3* and human *APOC3* (Fig 6M-P). There were no significant changes in the TG and TC metabolism-related genes in the liver, such as *Apoc2*, *Apoa5*, *Apoe*, *Apob*, *Mttp*, *Fasn*, *Srebp1c*, *Ldlr*, and *Pcsk9* (Supplementary Fig 10). *Angptl3* ASO administration did not significantly change body weight or systolic blood pressure (Fig 6Q-R). Overall, our findings demonstrate that administration of *Angptl3* ASO protected against hypertriglyceridemia accelerated AAA development in h*APOC3* Tg mice.

**Fig. 6:**
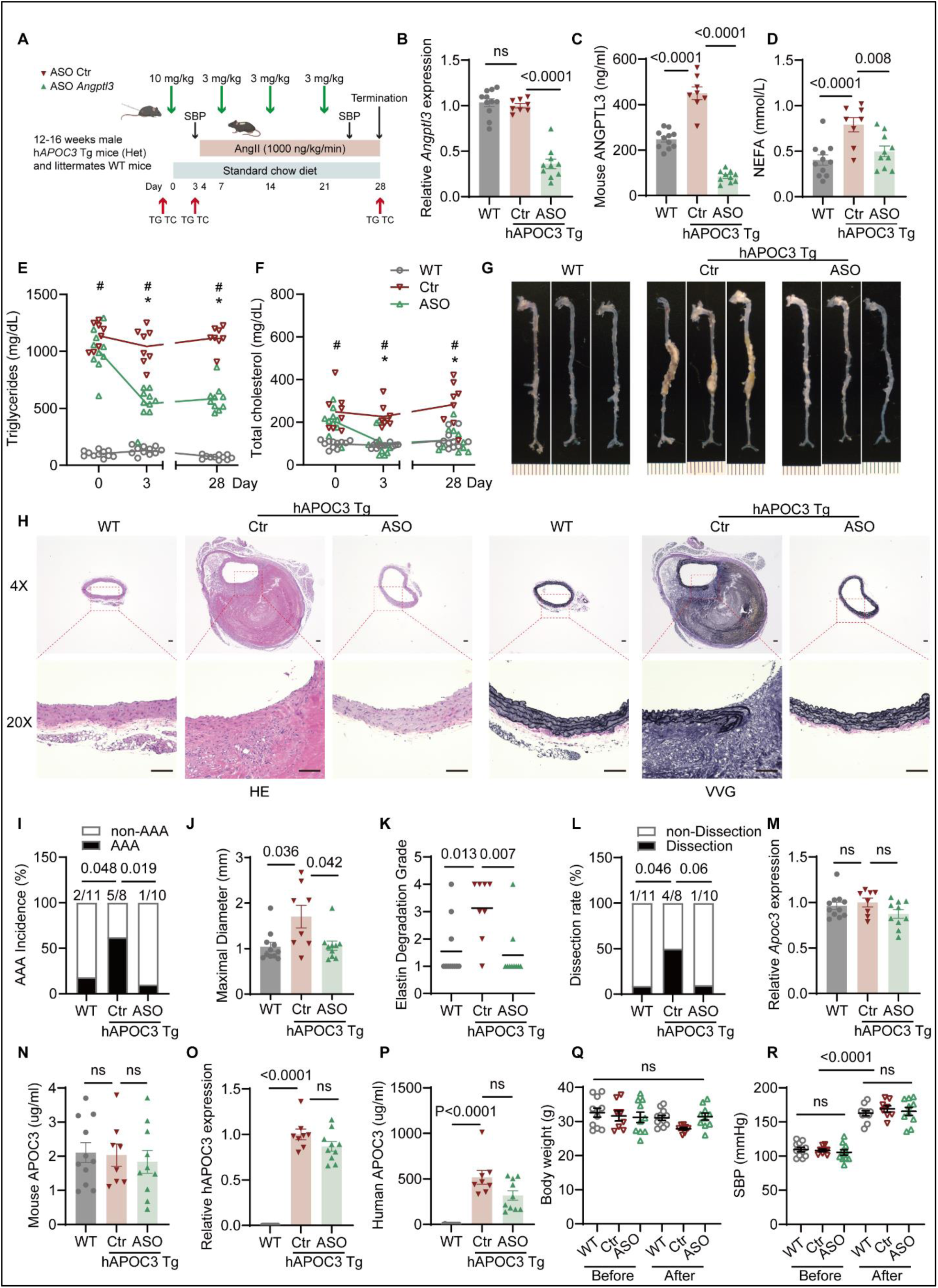
Administration of *Angptl3* ASO prevents AAA formation in h*APOC3* Tg mice. **A**, Design of the *Angptl3* ASO study in human *APOC3* Tg mice. Twelve- to 16-week-old male h*APOC3* Tg mice were given one injection of *Angptl3* ASO (10 mg/kg) or scrambled ASO by subcutaneous administration. After 3 days, mice were infused with AngII (1,000 ng/kg/min) for 25 days. Three more injections (3 mg/kg) were conducted on days 7, 14, and 21. A wild-type group was included, receiving injections of scramble ASO. At the end of the study, aortas, livers, and plasma were harvested. **B**, Relative abundance of *Angptl3* mRNA in livers of the three groups. **C-D**, Plasma ANGPTL3 protein concentrations (**C**) and non-esterified fatty acids (**D**) at the endpoint. **E** to **F**, Plasma triglycerides (TG) (**E**) and total cholesterol (TC) (**F**) concentrations on days 0, 3, and 28. **G**, Representative aortic trees from the 3 groups. **H**, Representative H&E staining (left) and Verhoeff-Van Gieson (VVG) staining (right) of suprarenal abdominal aorta sections. **I-L**, Quantifications of AAA incidence (**I**), maximal aorta diameter (**J**), elastic fiber degradation score (**K**), and dissection rate (**L**) among the 3 groups. **M**, Relative abundance of *Apoc3* in the liver. **N**, Plasma mouse-specific APOC3 concentrations were measured by ELISA at the endpoint. **O**, Relative liver abundance of human *APOC3*. **P**, Plasma human APOC3 concentrations were determined by ELISA at the endpoint. **Q**, Body weight comparisons among the 3 groups before and at the end of the study. **R**, Systolic blood pressure comparisons among the 3 groups before and at the end of the study. Data are presented as circles/dots and Mean ± SEM or Mean only. Statistical analyses were conducted as follows: One-way ANOVA followed by Sidak post hoc analysis for **B**, **C**, **D**, **E,** day 3 of **F, M, N, R**; Kruskal-Wallis test followed by Dunn’s post hoc analysis for day 0 and 28 of **F, J**, **K**, **O**, **P, Q**; Chi-Squared test for **I**, **L**. Scale bars: 1 mm in **G**, 100 µm in **H**. SBP, systolic blood pressure; NEFA, non-esterified fatty acids; #, *P* < 0.005 of the comparison of Ctr and WT group. *, *P* < 0.005 of the comparison of Ctr and ASO group.

### *Angptl3* ASO inhibits AAA development in *Apoe*-deficient mice

ApoE-deficient mice have high levels of VLDL due to impaired clearance of chylomicron and VLDL. Compared with age and sex-matched C57BL/6J mice, *Apoe*-deficient mice show significantly increased plasma levels of TC and moderately increased TG levels when fed a standard rodent diet.^38^ Similar to the observation in h*APOC3* Tg mice, administration of *Angptl3* ASO dramatically reduced hepatic *Angptl3* mRNA expression and the circulating levels of ANGPTL3 (Fig 7A-C). As expected, administration of the *Angptl3* ASO significantly decreased TG concentrations by 50%, NEFA by 31%, and slightly decreased TC concentrations by 8% in *Apoe*-deficient mice fed a standard rodent laboratory diet (Fig 7D-F). We also used size exclusion chromatography to separate lipoprotein classes and found decreased TG concentrations, mainly in VLDL and IDL (Fig 7I), while cholesterol concentrations were slightly reduced in VLDL and HDL (Fig 7J). Blood pressure measurements and body weight recordings revealed no significant difference between control ASO and *Angptl3* ASO-administered mice (Fig 7G, H). Consistent with the literature, AAA incidence in *Apoe*-deficient mice was 83% in the control group (Fig 7K, N). *Angptl3* ASO administration largely inhibited AAA development, as evidenced by significantly decreased AAA incidence and maximal aortic diameters (Fig 7N, O). Depletion of hepatic *Angptl3* had limited effects on the expression of genes involved in lipogenesis and cholesterol metabolism (Fig 7P). Taken together, these data demonstrated that lowering TG and NEFA concentrations by *Angptl3* ASO inhibited AAA development in *Apoe*-deficient mice.

**Fig. 7:**
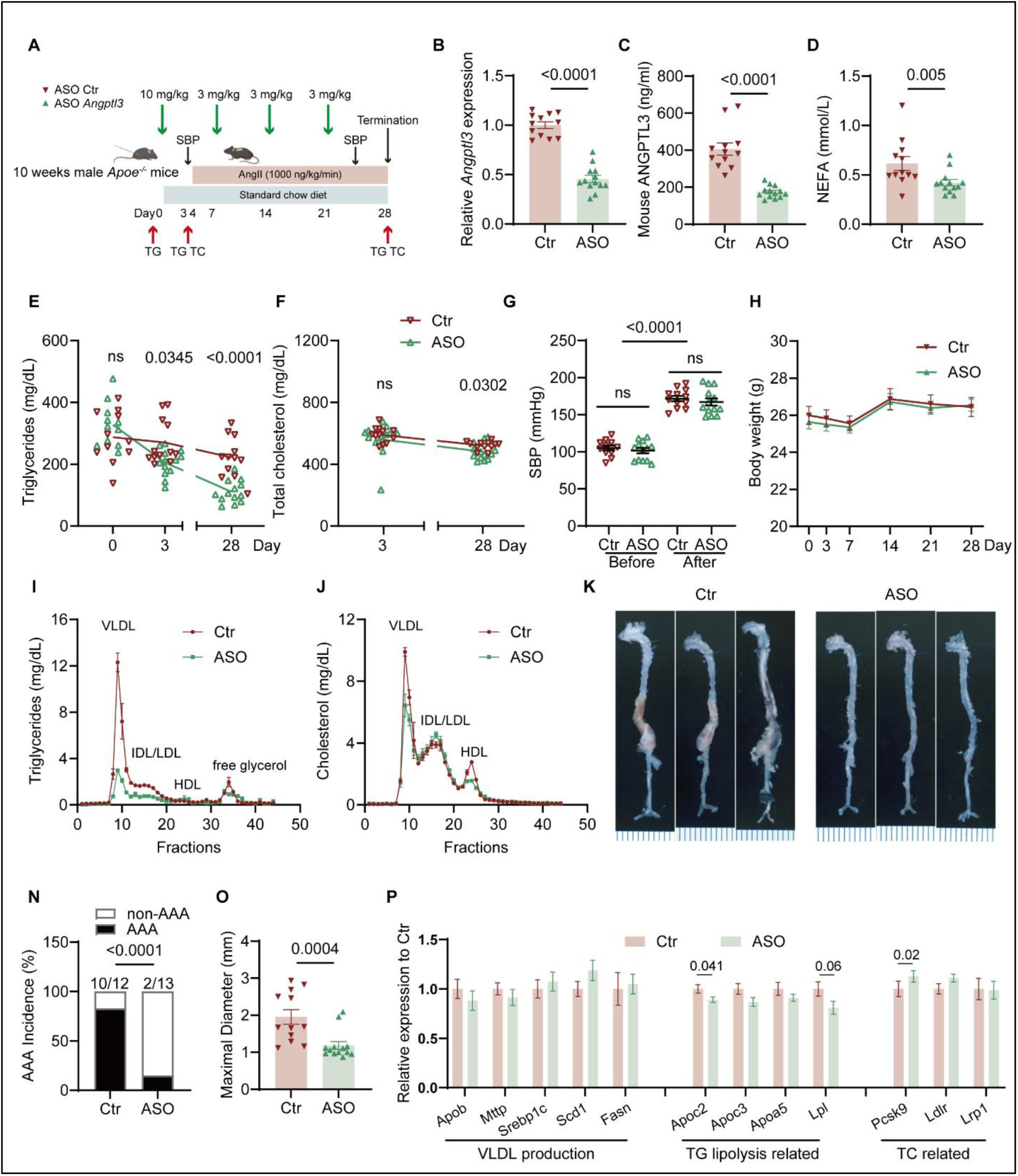
Administration of *Angptl3* ASO prevents AAA formation in *Apoe*-deficient mice. **A**, Design of the *Angptl3* ASO study in male *Apoe*-deficient mice. Ten-week-old male *Apoe*-deficient mice were given a subcutaneous injection of *Angptl3* ASO (10 mg/kg; n = 13) or scrambled ASO (n = 12). After 3 days, mice were infused with AngII (1,000 ng/kg/min) for 25 days. Three more injections were administered on days 7, 14, and 21. At the end of the study, aortas, livers, and plasma were harvested. **B**, Relative abundance of *Angptl3* mRNA in the liver. **C-D**, Plasma ANGPTL3 protein concentrations (**C**), and non-esterified fatty acids (**D**) at the endpoint. **E,** Plasma triglycerides (TG) levels on days 0, 3, and 28. **F**, Plasma total cholesterol (TC) concentrations on days 3 and 28. **G**, Systolic blood pressure before and after AngII infusion. **H**, Body weight changes. TG (**I**) and cholesterol (**J**) concentrations of size exclusion chromatography fractionated plasma from animals (n = 4 in the control group, n = 4 in the *Angptl3* ASO group) determined by enzymatic assays. Fractions 8 to 11 contained VLDL, fractions 12 to 17 contained IDL and LDL, and fractions 22 to 25 contained HDL. **K**, Representative aortic images from the 2 groups. **N-O**, Quantification of AAA incidence (**N**) and maximal aortic diameter (**O**). **P**, Relative liver mRNA abundance of genes related to VLDL production (*Apob, Mttp, Srebp1c, Scd, Fasn*), TG lipolysis (*Apoc2, Apoc3, Apoa5, Lpl*), and TC regulation (*Pcsk9, Ldlr, Lrp1*). Data are presented as circles/dots and/or Mean ± SEM. Statistical analyses were conducted as follows: Student’s t-test for **B**, **C**, day 0 and 28 of **E**, day 28 of **F**; Mann-Whitney U test for **D**, day 3 of **E** and **F**, **O**; One-way ANOVA followed by Sidak post hoc analysis for **G**; Chi-Squared test for **N;** Student’s t-test or Mann-Whitney U test for **P**. Scale bars: 1 mm in **K**. SBP, systolic blood pressure; NEFA, non-esterified fatty acids.

## Discussion

AAA screening programs have identified many asymptomatic AAA and effectively reduced aneurysm-related mortality. Thus, limiting aneurysm growth and reducing the risk of aneurysm rupture has become a key priority for the treatment of AAA.^39^ Based on the genetic, proteomic, and metabolomic findings, we identify that increased TG concentrations accelerate AAA formation, dissection, and rupture using three hypertriglyceridemic mouse models. Of clinical relevance, we provide evidence that lowering TG and NEFA concentrations by *Angptl3* ASO administration dramatically inhibited AAA development and dissection.

Mendelian Randomization (MR) using protein quantitative trait loci (pQTLs) as instrumental variables (IVs) can help infer causality and identify drug targets for complex diseases. By integrating pQTLs from ten large GWAS on circulating proteins, we updated the ‘actionable’ druggable target pool into an enlarged set of 2,698 proteins. Genetically determined APOA5 and LPL, two critical molecules regulating triglyceride metabolism, were negatively associated with AAA risk. In contrast, the variants related to APOC3 with increased TG concentrations were associated with increased AAA risk, underscoring the role of triglyceride metabolism in human AAA development, consistent with other genetic findings.^7, 8^ We further explored the causal effects of metabolites on AAA risk using IVs from 233 NMR-measured circulating metabolites.^20^ The strong associations found in VLDL and its subfractions as well as free fatty acid concentrations, further highlighted the importance of TG metabolism in AAA. Collectively, these findings emphasize the significance of TG in AAA risk and provide potential candidates for AAA prevention and treatment.

According to the literature and our experience, young C57BL/6J mice, when fed a standard rodent laboratory diet, have low AAA incidence when infused with AngII.^26, 40^ Hyperlipidemia increases the AAA incidence during AngII infusion in *Ldlr* and *Apoe*-deficient mice or C57BL/6J mice with an AAV expressing a gain-of-function mutant of PCSK9, in which *Ldlr*-deficient mice and PCSK9 overexpressing mice are fed a Western diet to augment hypercholesterolemia.^26^ Compared with C57BL/6J mice, *Ldlr* or *Apoe*-deficient mice have significantly increased TC and TG concentrations, which mainly distribute in VLDL fractions using size exclusion chromatography to separate lipoprotein classes,^41^ indicating that increased concentrations of large, TG-rich lipoproteins are associated with AAA formation. Our previous study demonstrated that AAA development is accelerated with modestly hypercholesterolemic conditions, but further elevation of cholesterol above a threshold level does not enhance AAA progression.^40^ Here, we present that increased TG concentrations contribute to AAA development, dissection, and rupture in a TG-concentration-dependent manner (Supplementary Table 7) using three hypertriglyceridemia mouse models. The moderately increased TG concentrations in *Apoa5* deficiency mice accelerated AAA development, dramatically increased TG concentrations in h*APOC3* transgenic mice caused AAA dissection, and severely high TG concentrations resulted in aortic rupture and death in inducible *Lpl*-deficient mice. We also evaluated the effects of increased TG concentrations on aneurysm growth in the elastase-induced AAA model. Previous studies have demonstrated that the aneurysm growth in the PPE model is independent of hypercholesterolemic conditions using C57BL/6J mice with PCSK9 overexpression or *Apoe*-deficient mice.^27, 42^ Using the PPE-induced AAA model, we demonstrated that increased TG concentrations aggravated AAA growth in both male and female h*APOC3* Tg mice. Furthermore, clinical observation shows that plasma TG concentrations are higher in AAA patients than in controls.^9^ In a prospective epidemiological study (the British United Provident Association [BUPA] study), the risk of death from AAA rupture is strongly related to serum TG concentrations.^10^ Together, these findings pave the way for future studies of the role of triglyceride and related metabolites in AAA development.

Importantly, our study demonstrates that lowering TG and NEFA levels by administrating *Angptl3* ASO inhibits AAA development in both h*APOC3* transgenic mice and *Apoe*-deficient mice. ApoC-III raises TG concentrations by inhibiting LPL activity, reducing hepatic uptake of TG-rich lipoproteins, and promoting VLDL secretion. ApoE acts as a critical ligand for the LDL receptor and the LDL receptor-related protein, facilitating the uptake of chylomicron and VLDL remnants into hepatocytes. Deficiency of ApoE causes elevated TG and TC levels. In both *APOC3* transgenic mice and *Apoe*-deficient mice, triglycerides and cholesterol are mainly distributed in VLDL fractions. Administration of *Angptl3* ASO dramatically reduced TG levels in VLDL and IDL fractions, consistent with decreased TG concentrations in plasma, supporting the hypothesis that large-sized TG-rich lipoproteins and related metabolites contribute to AAA progression and rupture. These findings also provide evidence that lowering TG-rich lipoproteins is a potential therapeutic strategy. ANGPTL8 and ANGPTL3 form a complex that markedly inhibits LPL activity. Similar to the effects of *Angptl3* ASO on AAA, *Angplt8*-deficiency or knockdown reduced AAA formation by lowering TG concentrations and suppressing the inflammatory response in *Apoe*-deficient mice.^43^ Administration of fenofibrate also reduced the severity of experimental AAA in *Ldlr*- and *Apoe*-deficient mice.^44, 45^ However, fenofibrate did not significantly limit AAA growth in AAA patients in FAME-2 (Fenofibrate in the Management of Abdominal Aortic Aneurysm 2), a placebo-controlled clinical trial.^46^ We should note that it may take an estimated five years for a small AAA with a diameter of 40–55 mm to reach 55 mm, the size threshold for intervention.^39^ In the FAME-2 study, AAA diameters in the control group increased only about 1 mm after a 24-week observation period, making it hard to determine the effects of fenofibrate. Additionally, AAA diameters in the two groups began to diverge at the end of the evaluation, suggesting that a long-term clinical trial with a larger sample size is needed to carefully assess the effects of triglyceride-lowering on AAA growth and rupture. Statins primarily target TC but also have a moderate TG-lowering effect (10–30%), depending on the statin type and dose. While observational studies have found potential benefits of statins in AAA management by reducing inflammation, stabilizing the aortic wall, and lowering TC and TG levels, their impact on AAA growth and rupture risk remains controversial and inconclusive^47–49^. This variability may be influenced by differences in statin types, dosages, and genetic metabolic-related factors. Therefore, high-quality randomized controlled trials are needed further to clarify their role in AAA prevention and treatment.

AAA is characterized by pathological remodeling of the aortic ECM, including elastolysis and collagenolysis. LOX is a crucial enzyme involved in the cross-linking of collagen and elastin in the ECM to maintain the structural integrity of the aortic wall. Impairments in LOX activity due to genetic mutations can lead to connective tissue disorders and promote aorta dissection.^50^ *Lox*-deficient mice exhibit severe defects in vascular development, leading to perinatal death from aortic aneurysm.^32^ β-Aminopropionitrile (BAPN) is a well-known inhibitor of LOX and has been used extensively in research to accelerate AAA formation and rupture.^51^ LOX is initially synthesized as a preproprotein and secreted into the extracellular space, where the prolysyl oxidase is cleaved by proteases, including BMP-1, ADAMTS-2, and ADAMTS-14. Increased TG concentrations or the presence of palmitate dramatically blocked LOX maturation in aortas and HASMC. Based on RNA-seq findings, *BMP1* and *ADAMTS2* are highly abundant in HASMC. PA downregulated the expression of *BMP1* and *ADAMTS2*, causing decreased LOX maturation. In this study, we confirmed BMP-1 is the predominant enzyme regulating LOX maturation in HASMC. The increase in LOX transcription upon *BMP1* knockdown or palmitate treatment was likely a compensatory response; however, it was insufficient to compensate for the loss of mature LOX. Increasing the substrate availability through LOX overexpression could partially compensate for reduced BMP-1 function and produce a certain amount of mature LOX. Further analysis of liver-aorta crosstalk using data from the GTEx project revealed that the liver is predicted to influence aortic gene expression through the secretion of APOC3. Notably, higher liver *APOC3* levels are associated with lower *BMP1* expression in the aorta. Previous studies have shown that LOX overexpression can inhibit AAA development in the CaCl₂-induced AAA model.^52^ In our AngII model in h*APOC3* Tg mice, local LOX overexpression in the suprarenal aorta completely abolished the effects of high TG on promoting suprarenal AAA. Additionally, PA induces vascular SMC apoptosis, promotes inflammatory responses, enhances oxidative stress, and triggers proliferation and migration, contributing to the progression of vascular diseases such as atherosclerosis and hypertension.^53^ In this study, we demonstrated that PA, as a metabolite of plasma triglycerides or endogenously synthesized in the liver, may contribute to AAA development by inhibiting LOX maturation. These findings uncover a novel liver-aorta regulatory mechanism where APOC3-induced hypertriglyceridemia suppresses BMP-1 and impairs LOX maturation, potentially driving AAA formation.

Plasma TG concentrations include the content of various lipoprotein particles. Multiple genes, such as LPL, APOC3, APOC2, APOA5, and ANGPTL3, influence TG metabolism and regulate plasma TG concentrations. Among them, APOC3 and ANGPTL3 are targets for drug development because loss-of-function mutations in APOC3 and ANGPTL3 are associated with improved lipid profiles and reduced cardiovascular risk. Preclinical and clinical data have well documented that ApoC-III or ANGPTL3 inhibitors effectively reduce severe hypertriglyceridemia. These inhibitors include antisense nucleotides (ASO),^14, 16, 37^ monoclonal antibodies (mAb),^54^ small interfering RNA (siRNA),^55, 56^, and CRISPR-cas gene editing strategies.^57, 58^ Preclinical and clinical data have demonstrated that ANGPTL3 inhibitors are highly effective for treating severe hypertriglyceridemia by upregulating LPL activity and facilitating the hydrolysis and clearance of TG-rich lipoproteins. As h*APOC3* transgenic mice have very high APOC3 and TG concentrations, *Angptl3* ASO was administrated at 50 mg/kg/week in previous studies.^16^ In this study, administration of an *Angptl3* ASO at a much lower dosage effectively reduced AAA development in h*APOC3* transgenic mice and *Apoe*-deficient mice through reducing TG concentrations, especially TG concentrations in VLDL fractions. We observed that the administration of *Angptl3* ASO mainly decreased plasma TG concentrations with a limited reduction in plasma TC concentrations, further supporting the idea that managing TG-rich lipoprotein concentrations is a potential therapeutic strategy for treating AAA.

This study has several limitations. First, while our animal models exhibit increased TG levels, some also show concurrent alterations in TC levels, making it challenging to fully distinguish their independent effects. However, the strong correlation between TG levels and AAA severity, the mechanistic link between TG and LOX inhibition, and the therapeutic efficacy of TG-lowering strategies via *Angptl3* ASO administration collectively support a direct role of hypertriglyceridemia in AAA pathogenesis. Second, although the *Lpl*-KO mouse model provides valuable insights into the impact of severe TG elevation on AAA, its extreme hypertriglyceridemia limits translational relevance. To address this, we incorporated moderate hypertriglyceridemia models for mechanistic and therapeutic studies. *Lpl*-KO mice exhibit severely elevated TG and chylomicron levels, closely mirroring human familial chylomicronemia syndrome, a rare and severe condition affecting approximately 1 in 1,000,000 individuals^59^. In contrast, heterozygous h*APOC3*-Tg, *ApoA5*-deficient, and *ApoE*-deficient mice develop mild to moderate hypertriglyceridemia when fed a standard chow diet, resembling familial hypertriglyceridemia, a more common disorder affecting approximately 1 in 100 individuals.^60^ Future studies utilizing animal models with selective modulation of TG, cholesterol, or TG-rich lipoprotein levels will be essential to clarify further their distinct contributions to AAA pathogenesis and therapeutic targeting.

In summary, the present studies, combined with recent genetic, proteomic, and metabolomic findings, indicate that increased TG concentrations contribute to an increased risk of AAA. Individuals with higher genetic predisposition for AAA may benefit from targeted interventions. Understanding the association between triglyceride concentrations and AAA risk has important clinical implications. Notably, our results demonstrate that ASO therapy targeting liver ANGPTL3 holds potential as a therapeutic approach to reduce AAA risk. These findings underscore the need for additional experimental and clinical investigations into the mechanisms and therapeutic strategies for AAA treatment through triglyceride-lowering therapies.

## Data availability

All genetic data used in this study were obtained from publicly available consortiums. The RNAseq original data generated during this study will be deposited in a publicly accessible database upon acceptance of the manuscript.

