## Supplementary Materials for "Hypertriglyceridemia as a Key Contributor to Abdominal Aortic Aneurysm Development and Rupture: Insights from Genetic and Experimental Models"

### Methods

#### Causal effects of circulating proteins on AAA risk

Genetic variants that affect protein concentrations in a 'cis' manner can serve as valuable tools for guiding therapeutic targeting as they mimic the beneficial or harmful effects observed by pharmacological modification[1, 2]. Our group previously utilized 717 cis-pQTLs compiled by Zheng. et al.[3] from 5 pQTL studies and conducted Mendelian Randomization (MR) analyses to assess their causal effects on AAA risk[4]. Five later published large-scale genome-wide association studies (GWAS) of plasma proteins were incorporated to broaden the investigation[5] in the present study. Briefly, the cis-pQTLs of circulating proteins from a total of 10 genome-proteome–wide association studies were retrieved, including tier1 cis instrument variables reported by Zheng et.al[6-11], as well as sentinel cis-pQTLs from five subsequent publications[12-16]. Sex chromosome variants and variants with minor allele frequency of less than 0.01 were excluded. Next, associations of the selected variants with AAA were retrieved from our previous large-scale GWAS meta-analysis involving 39,221 individuals with AAA and over 1 million controls[4]. Data on exposure and outcome were then harmonized to ensure that the effect of an SNP on exposure and the outcome corresponded with the same allele. After standardizing protein target names to Ensemble gene ID, variant-protein pairs from different studies were integrated accordingly. For proteins reported by Zheng et al., the same cis-pQTLs were employed. For proteins reported only in later studies, multiple statistics could exist for the same variant-protein pair due to factors such as multiple probes or different protein isoforms. In these cases, we selected the statistic with the highest F-statistics, an indication of instrument strength[2]. In the same way, the variant with the highest F value for proteins with multiple available variants was selected. Ultimately, 2,698 circulating proteins were assigned unique and optimal instrumental variables. These were used in two-sample MR analyses to assess causal effects on AAA using the MR-Wald ratio method[17]. The false discovery rate was controlled by the Bonferroni correction.

We also applied multi-instrument variable (multi-IV) MR for the circulating proteins that passed multiple tests. We retrieved the available summary statistics for each of the proteins from 3 pQTL studies[12, 15, 16], which have the largest sample sizes and protein profile numbers. We selected the instrumental variables (IVs) and performed the MR analysis separately based on the summary statistics from each study. Briefly, IVs were selected by retrieving independent genetic variants ($r2<0.001$, kb = 10,000, based on individuals with European ancestors from the 1000 Genomes Project) of the corresponding protein targets at a genome-wide significance level ($P<5\times10^{-8}$). Three MR methods, including inverse variance weighted (IVW) MR, weighted median-based regression, and MR-Egger analysis, were then applied. The inverse variance–weighted (IVW) MR[18] provides the highest precision while assuming that all SNPs are valid instrumental variables[19]. It offers an unbiased estimate when no horizontal pleiotropy is present or when horizontal pleiotropy is balanced. To account for potential pleiotropy, we also applied weighted median-based regression and MR-Egger analysis. The weighted median estimates are almost as precise as IVW estimates, but they require that at least half of the MR instrument weights on the exposure be valid[19]. The MR-Egger analysis, though having lower precision, can detect and correct for pleiotropy, allowing for causal inference even if all genetic variants have pleiotropic effects. A consistent effect across all three methods is unlikely to be a false positive, resulting in increased robustness.

**Prioritizing the role of major lipids and lipoproteins as risk factors for AAA**

To prioritize the role of circulating lipids and lipoproteins in AAA risk, we applied Mendelian Randomization Bayesian Model Averaging (MR-BMA) analyses[20] by jointly considering 5 correlated exposures including low-density lipoprotein cholesterol (LDL-C), high-density lipoprotein cholesterol (HDL-C), apolipoprotein A1 (ApoA1), apolipoprotein B (ApoB), and triglycerides (TG). After retrieving independent genetic variants ($r2<0.001$, kb = 10,000, based on individuals with European ancestors from the 1000 Genomes Project) associated with any major lipoprotein-related trait (total cholesterol, LDL-C, HDL-C, or TG) at a genome-wide significance level ($P<5\times10^{-8}$) in the Global Lipids Genetics Consortium GWAS (2013)[21], we removed influential variants based on the Cook's distance and outliers based on the q-statistic. The genetic associations of the selected variants with LDL-C, HDL-C, ApoA1, ApoB, and TG were from the UK biobank study[22], and their association with AAA was from our previous GWAS analysis[4]. Full details of the MR-BMA methodology can be found elsewhere[20]. Briefly, MR-BMA evaluates multiple potential causal models incorporating various subsets of exposures. For each exposure, a Marginal Inclusion Probability (MIP) is computed, indicating the likelihood of a metabolite being included in the true causal model across iterations (z). We used the following parameters: z = 1,000 iterations, prior probability set to 0.1, and prior variance ($\sigma$^2) to 0.25. An empirical permutation procedure was used to calculate P-values, which were adjusted for multiple tests using the Benjamini-Hochberg false discovery rate (FDR) procedure.

**Causal effects of circulating metabolites on AAA risk**

The genetic instruments for circulating metabolites were retrieved from a recently published study[23], which conducted GWAS analysis on 233 NMR-measured metabolites. The inverse variance weighted (IVW) MR was used as the primary method[18], which provided the highest precision while assuming that all SNPs are valid instrumental variables. To address potential pleiotropy, MR-Egger analysis and weighted median-based regression were performed as sensitivity analyses[19]. Consistent findings across all three methods bolstered robustness and minimized the risk of false positives. For illustration, the diameters of 14 lipoprotein subclass particles are plotted based on mean values from 5,651 participants in the Northern Finland Birth Cohort 1966 (NFBC66)[24].

### Causal effects of plasma triglycerides levels on circulating fatty acid concentrations

Genetic instruments and their associations with plasma TG were determined by retrieving independent genetic variants (r2<0.001, kb = 10,000, based on individuals with European ancestors from the 1000 Genomes Project) associated with TG at a genome-wide significance level ($P<5\times10^{-8}$) in the Global Lipids Genetics Consortium GWAS (2013)[21]. Their associations with circulating total fatty acid level were retrieved from the NMR-based GWAS[23]. Their associations with circulating levels of palmitic acid (16:0), stearic acid (18:0), palmitoleic acid (16:1n-7), and oleic acid (18:1n-9) were retrieved from the Cohorts for Heart and Aging Research in Genomic Epidemiology (CHARGE) Consortium[25]. The inverse variance weighted (IVW) MR was used as the primary method[18], which provided the highest precision while assuming that all SNPs are valid instrumental variables. To address potential pleiotropy, MR-Egger analysis and weighted median-based regression were performed as sensitivity analyses.

### Materials and reagents

Antibody against LOX (Cat: A11504) was purchased from Abclonal. Antibodies against β-actin (Cat: 3700) and GFP (Cat: 2956) was purchased from Cell Signaling Technology (CST, Danvers, MA). Angiotensin II (AngII, Cat: 4006473) was purchased from Bacham. Bovine serum albumin (BSA, Cat: A7030-100G) was purchased from Sigma. Palmitic acid was purchased from Sigma (Cat: P0500). BMP1 siRNA (Assay ID: 105352), ADAMTS2 siRNA (Assay ID: 105359), ADAMTS14 siRNA (Assay ID: 105394), and siRNA control were obtained from Invitrogen.

### Antisense Oligonucleotides Directed to Murine Angptl3[26]

For the ASO-mediated TG lowering murine studies, chimeric 20-mer phosphorothioate antisense oligonucleotides (ASOs) directed to murine *Angptl3* mRNA (5'-GACATGTTCTTCACCTCCTC-3' or control ASO (5'-CCTTCCCTGAAGGTTCCTCC-3') were produced by BOC Sciences. The ASOs contain 2'-O-methoxyethyl (2'-MOE) groups at positions 1-5 and 16-20 and have been modified by the addition of a covalently bonded triantennary N-acetyl galactosamine (GalNAc).

### Animal preparation

Breeding pairs of *Lpl* floxed mice without or with the β-actin driven tamoxifen-inducible MerCreMer transgene in C57BL/6J background were provided by Dr. Ira Goldberg[27]. Male MerCreMer +/0 *Lpl* $^{f/f}$ mice and female MerCreMer 0/0 *Lpl* $^{f/f}$ mice were bred to generate offspring.

C57BL/6J mice (Stock No: 000664), human *APOC3* transgenic (h*APOC3* Tg, Stock No: 006907) mice, and *Apoe*-deficient mice (Stock No: 002052) were purchased from The Jackson Laboratory. *Apoa5*-deficient mouse was purchased from MMRRC (#011467-UCD; Davis, CA), and was backcrossed with C57BL/6J inbred mice for at least 8 generations. h*APOC3* heterozygous mice were bred with C57BL/6J mice to generate h*APOC3* heterozygous and littermate h*APOC3* wild-type control mice. For studies involving *Apoe*-deficient mice or C57BL/6J mice, animals were purchased from The Jackson Laboratory and acclimatized for at least 1 week at the University of Michigan before any procedures were initiated. All animal procedures were conducted in accordance with protocols approved by the University of Michigan Institutional Animal Care & Use Committee.

**Induction of abdominal aortic aneurysm**

AAA is defined as a ≥ 50% increase in suprarenal (AngII-induced model) and infrarenal (PPE model) abdominal aortic diameter. In the AngII model, we defined dissection in the suprarenal aortic region as a transmural break of the media layers that leads to exit of blood to provoke adventitial dissection[28], which is characterized by the presence of vascular hematoma or remodeled thrombi that are visible during tissue harvesting or sectioning. Aortic rupture is defined as death resulting from aortic rupture (either thoracic or abdominal) during AngII infusion. Most of the animal models used in the present study utilized AngII infusion, as described earlier[29], in which animals were infused subcutaneously via a minipump (Alzet, model 2004 or model 2002). Tail-cuff-based systolic blood pressure measurements were applied before pump implantation and before tissue harvesting.

In MerCreMer 0/0 *Lpl* f/f (wild-type littermate controls) mice and MerCreMer +/0 *Lpl* f/f (inducible *Lpl*-deficient) mice fed standard rodent laboratory diet (TD.2918), tamoxifen (75 mg/kg/day) was injected intraperitoneally for 5 consecutive days at the age of 7-10 weeks. Two weeks after the start of tamoxifen injections, mice were fed a low-cholesterol, Western diet (TD.05311, Harlan Teklad, Madison, WI) for 1 week before osmotic pumps were implanted. Western diet feeding was continued during AngII infusion for 4 weeks. Aortic aneurysm is defined as aortic rupture during the study or a ≥ 50% increase in suprarenal abdominal aortic diameter.

In the *Apoa5* knockout experiment, 10-12-week-old *Apoa5*-deficient mice and the littermate controls were fed on a standard rodent laboratory diet (LabDiet, 5L0D) and infused subcutaneously with AngII (1,500 ng/kg/min) for 4 weeks to induce AAA. In an independent cohort, we infused the male mice with AngII (1,000 ng/kg/min) for 2 weeks. The plasma was collected to measure the total cholesterol (TC), TG, non-esterified fatty acids and was used to run size exclusion chromatography.

In the h*APOC3* overexpression experiment, 12-16-week-old h*APOC3* Tg (heterozygous) mice and littermate controls were fed on a standard rodent laboratory diet (LabDiet, 5L0D) and infused subcutaneously with AngII (1,000 ng/kg/min) for 4 weeks to induce AAA.

For investigating the abundance of mature LOX in the abdominal suprarenal aorta at the initial stage of AAA formation, 12-16-week-old male h*APOC3* Tg (heterozygous) mice and littermate controls were infused subcutaneously with AngII (1,000 ng/kg/min) or saline for 7 days.

For the local LOX overexpression experiment, a high-concentration F-127 gel solution was prepared by dissolving 1 g F-127 in 2 mL PBS overnight and stored at 4°C. 12-16-week-old male h*APOC3* Tg (heterozygous) mice and littermate controls were anesthetized via intraperitoneal injection of a ketamine (90 mg/kg) and xylazine (5 mg/kg) mixture. The perivascular adipose tissue was carefully detached from the suprarenal abdominal aorta. Next, $9 \times 10^8$ pfu of adenovirus expressing GFP or LOX were mixed with the prepared F-127 gel solution at a 1:1 ratio (v:v) and injected immediately into the region between the suprarenal abdominal aorta and the surrounding adipose tissue. After a 20-minute incubation period, the abdominal cavity was closed, and the mice were implanted with AngII (1,000 ng/kg/min) for 18 days to induce AAA. All experiments were conducted using mice fed a standard rodent laboratory diet (LabDiet, 5L0D).

In a prevention study using *Angptl3* ASO in the h*APOC3* Tg mice, 12-16-week-old male h*APOC3* Tg (heterozygous) were subcutaneously injected with *Angptl3* ASO (10 mg/kg) or control ASO. After 3 days, the mice were infused subcutaneously with AngII (1,000 ng/kg/min) for 25 days to induce AAA, and additional ASO injections were performed on days 7, 14, and 21, with the dose decreased to 3 mg/kg. A littermate WT group was also included in the control ASO injection. All experiments were performed in mice with a standard rodent laboratory diet (LabDiet, 5L0D).

In a prevention study using *Angptl3* ASO in *Apoe*-deficient mice, 10-week-old male *Apoe*-deficient mice purchased from The Jackson Laboratory were subcutaneously injected with *Angptl3* ASO (10 mg/kg) or control ASO for 3 days, and infused subcutaneously with AngII (1,000 ng/kg/min) for 25 days to induce AAA and additional ASO injections were performed on day 7, 14, and 21, with the dose decreased to 3 mg/kg. All animals were fed a standard rodent laboratory diet (LabDiet, 5L0D).

The peri-adventitial elastase application-induced (PPE-induced) AAA model was performed as we described previously[30, 31]. For the PPE-induced AAA model in h*APOC3* Tg mice, 8-week-old Tg (heterozygous) mice and their littermate controls were anesthetized via intraperitoneal injection of a ketamine (90 mg/kg) and xylazine (5 mg/kg) mixture. The infrarenal abdominal aorta was isolated and surrounded by sterile gauze soaked in 30 µL of elastase (41 U/ml, Sigma, E1250). After a 30-minute incubation period, the gauze was removed, and the abdominal cavity was washed once with sterile saline before suturing. Mice were harvested 14 days after the PPE exposure.

Unless specified, no fasting procedures were conducted prior to sample collection. Mice were euthanized with $CO_2$ and infused with 8-10 ml of sterile saline. The suprarenal abdominal aorta (in the AngII model) or infrarenal abdominal aorta (in the PPE model) was isolated and measured promptly. The maximum abdominal aortic diameter was measured using a digital caliper by at least two individuals who were blinded to the experimental groups, and the mean value was used as the result.

### Histology analysis

At termination, the suprarenal abdominal aorta (in the AngII model) was carefully dissected, fixed in neutral buffered formalin (10% v/v), embedded in paraffin, and sectioned into 5 μm thick slices. The H&E, Masson's Trichrome, and Verhoeff-van Gieson (VVG) stainings were performed by the University of Michigan ULAM Pathology Core. Elastic fiber degradation was graded as follows: 1, <25% degradation; 2, 25-50% degradation;3, for 50-75% degradation; and 4, for >75% degradation, or dissection[31].

### Measurement of plasma lipid, human APOC3, mouse APOC3, mouse ANGPTL3 level, and free fatty acid

Whole blood was collected either from the facial vein during the experiment or through the heart at the time of tissue harvesting into EDTA-containing anticoagulant blood collection tubes without prior fasting unless specified. The blood was then centrifuged at 4°C at 1,500 g for 20 minutes, and the supernatant plasma was collected and either measured or immediately stored at -80°C.

Plasma total cholesterol (TC), TG, and non-esterified fatty acids (NEFA, or free fatty acids) concentrations were measured using enzymatic kits (FUJIFILM Wako Diagnostics). Samples with severe hemolysis were excluded from NEFA measurement. Plasma APOC3 concentrations were determined using species-specific ELISA kits for human APOC3 protein (Cat: ab154131, Abcam) and mouse APOC3 protein (Cat: LS-F22274, LS Bioscience). Plasma mouse ANGPTL3 levels were determined using the ANGPTL3 ELISA kit (Cat: MANL30, R&D Systems).

### Untargeted metabolomics in h*APOC3* Tg mice

Whole blood was collected from 14- to 18-week-old male h*APOC*3 Tg (heterozygous) mice and their littermate controls via cardiac puncture at the time of tissue harvesting. Blood was drawn into heparin-containing anticoagulant collection tubes and then centrifuged at 4°C at 1,500 g for 20 minutes. The supernatant plasma was collected and immediately stored at -80°C. Untargeted metabolomics analyses were performed by the Metabolomics Core at the University of Michigan. All animals were fed a standard rodent laboratory diet (LabDiet, 5L0D).

### Size Exclusion Chromatographic Resolution of Lipoproteins

Aliquots of plasma from 2 animals in the same group were pooled at a 1:1 ratio (*Apoe* ASO study) and centrifuged at 4°C, 10,000 g for 10 min. Aliquots of plasma from each animal in the *Apoa5* KO study were centrifuged at 4°C, 10,000 g for 10 min. The supernatant was then analyzed using a Waters HPLC system equipped with a Superose 6, 10/300 GL column (GE Healthcare, Piscataway, NJ). Samples were eluted with PBS at a flow rate of 0.5 ml/min and monitored at 220 nm. After removing the first 8 minutes' fractions, 44 more fractions (500 ul per fraction) were collected. The TG and cholesterol contents in each fraction were measured using enzymatic kits (FUJIFILM Wako Diagnostics).

### HASMC culture and treatment

Human aortic smooth muscle cells (HASMCs, CC-2571) were purchased from Lonza (Walkersville, MD) and Sigma (C-2305) . HASMCs were cultured in SMC Growth Medium 2 (Promo Cell, Germany) at 37°C with 5% $CO_2$ in a humidified incubator. Cells were passaged at a 1:3 ratio, and passages 4 to 6 were used for experiments. Prior to incubation, HASMCs at 80-90% confluence were incubated in Opti-MEM™ Reduced Serum Medium for 24 hours. For the palmitic acid (PA) incubation study, a 10% BSA (10% wt/vol) solution was prepared by dissolving fatty acid free BSA (Sigma) in sterile saline and stored at 4°C. PA was dissolved in saline at a 50 mM stock concentration and stored at -20°C. For each experiment, PA was thawed at 70°C and dissolved in 10% BSA to 5 mM. This solution was then added to Opti-MEM serum-reduced medium to obtain a final concentration of 250 µM. The control group was mixed with saline instead of PA to maintain a consistent BSA concentration. After indicated hours of incubation, cells were harvested for RNA extraction and whole-cell protein extraction. The conditioned medium was centrifuged at 300 g at 4°C for 5 mins and stored immediately at -80°C.

### Preparation of adenovirus

The full-length human *LOX* cDNA encoding *LOX* was subcloned into the pCR8⁄GW⁄TOPO entry vector (Invitrogen). After sequencing, the LR recombination reaction was carried out between the entry clone pCR8⁄GW⁄TOPO/*LOX* and the destination vector pAd/CMV/V5-DEST according to the manufacturer’s protocol (Invitrogen). The Ad293 cells were transfected with PacI linearized recombinant adenoviruses for packaging. After amplification, the recombinant adenoviruses were purified by $CsCl_2$ density gradient ultracentrifugation. Adenovirus titration was performed using the Adeno-X qPCR Titration Kit (Clontech). AdLacZ or AdGFP was used as controls.

### In vivo palmitic acid incubation

Ethyl palmitate (Tokyo Chemical Industry) was dissolved with lecithin (1.6% wt/vol; Thermo Scientific, 413102500) and glycerol (3.3% vol/vol) in water to produce a mixture containing ethyl palmitate (600 mM), lecithin (1.2% wt/vol), and glycerol (2.5% vol/vol), as reported previously[32]. The lecithin-glycerol-water solution was used as the control. 8-week-old C57BL/6J mice fed a normal rodent laboratory diet were intraperitoneally administered either ethyl palmitate or vehicle daily for 5 consecutive days (600 mg/kg). Prior to tissue harvest, mice were fasted for 16 hours, and the last injection was conducted 6 hours before euthanasia. At the time of tissue harvesting, mice were euthanized with $CO_2$ and infused with 8-10 ml of sterile saline. The suprarenal abdominal aorta was isolated and snap-frozen for protein extraction.

### RNA extraction, RT-qPCR, and RNA sequencing analyses

*RNA extraction, reserve transcription, and qPCR*

Total RNA was extracted from the liver or cultured HASMCs using RNeasy Mini kit (74106, QIAGEN, Hilden, Germany). cDNA samples were synthesized using oligo(dT) primers and the SuperScript III First-Strand Synthesis System (18080051, Invitrogen). For qRT-PCR analysis, cDNA reverse transcribed from 40 ng RNA was used. Relative mRNA expression was determined using the $2^{-\Delta\Delta Ct}$ method, with *Ppia* being the internal control. Primer sequences used are listed in Supplementary Table 6.

*RNA sequencing and quantification*

About 1 μg of RNA extracted from cultured HASMCs was submitted to the Advanced Genomics Core at the University of Michigan for RNA-seq analysis. cDNA libraries were prepared using the Illumina NEBNext Ultra RNA Library Prep Kit, and sequencing was conducted on the NovaX 10B 300 cycle to generate 150-base pair paired-end reads. Cutadapt (v2.3) was employed to remove potential low-quality sequences and adapter remnants. The quality of the trimmed data was assessed using FastQC (v0.11.8), and Fastq Screen (v0.13.0) was utilized to screen for various types of contamination. For alignment, reads were mapped to the reference genome GRCh38 (ENSEMBL), using STAR v2.7.8a (Dobin et al., 2013) and assigned count estimates to genes with RSEM v1.3.3 (Li and Dewey, 2011), which provides count values and Transcripts Per Million (TPM) values.

*Bioinformatic analysis*

Differentially expressed gene analysis was conducted using the R DESeq2 package[33] (v1.40.1). Differentially expressed gene was defined as $|log2foldchange| > 1$ and a False Discovery Rate (FDR) < 0.05. Biological process annotations for gene sets were analyzed using the enrichPathway function from the R clusterProfiler package[34] (v4.8.1). Gene Set Enrichment Analysis (GSEA) analysis was conducted using the GSEA function from the R clusterProfiler package[34] utilizing the ranking list of all genes sorted by fold change.

### Protein extraction and Western blotting of tissue and cells

Total protein from tissues or cells was extracted using RIPA buffer, separated by SDS-PAGE, and transferred onto nitrocellulose membranes. Membranes were blocked in TBST (Tris-buffered saline with Tween-20) containing nonfat dry milk (5% wt/vol) at room temperature for 30 minutes and then incubated with primary antibodies (β-actin, LOX, GFP) at 4°C overnight. After three washes with TBST, membranes were incubated with secondary antibodies (Li-Cor Biosciences, Lincoln, NE) at a 1:10,000 dilution for 30 minutes at room temperature. The membranes were then washed 3 more times with

TBST and scanned using the Odyssey Imaging System (Li-Cor Biosciences, Lincoln, NE). Band intensities were quantified using the LI-COR Image Studio Software.

**Western blotting of conditioned medium**

30 µL of conditioned medium was mixed with 10 µL of 4X SDS-loading buffer and boiled at 95°C for 5 mins, then used for Western blotting as described above. Whole-cell DNA was extracted using the Proteinase K digestion-based method, and the dsDNA concentration was used for normalization.

### Supplementary Figures

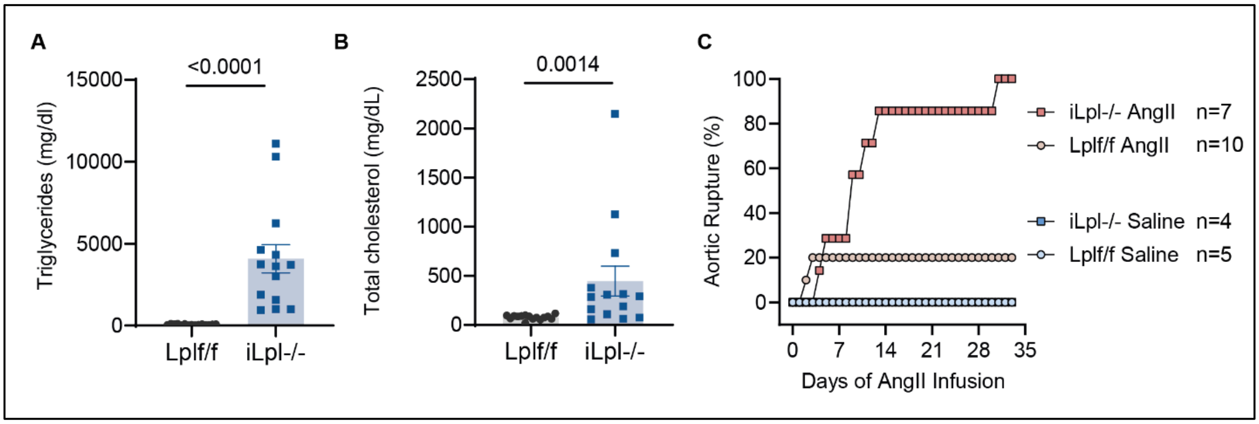


**Supplementary Fig. 1: Lpl inducible knockout promotes aortic rupture in male mice (replication study).**

Plasma triglycerides (**A**) and cholesterol (**B**) in *Lpl*f/f and i*Lpl*-/- male mice after 1 week of low cholesterol, Western diet feeding but before minipump implantation. Blood was collected from mice following a 4-hour fast. **C**, Aortic rupture incidence curve during 33-days AngII or saline infusion. Aortic rupture occurred for all i*Lpl*$^{-/-}$ mice in the AngII group (7/7) and for 2 *Lpl*$^{f/f}$ mice in the AngII group (2/10). No mice died in the saline-infused *Lpl*$^{f/f}$ (0/5) and i*Lpl*$^{-/-}$ (0/4) groups. Student’s t-test for **A**; Mann-Whitney U test for **B**.

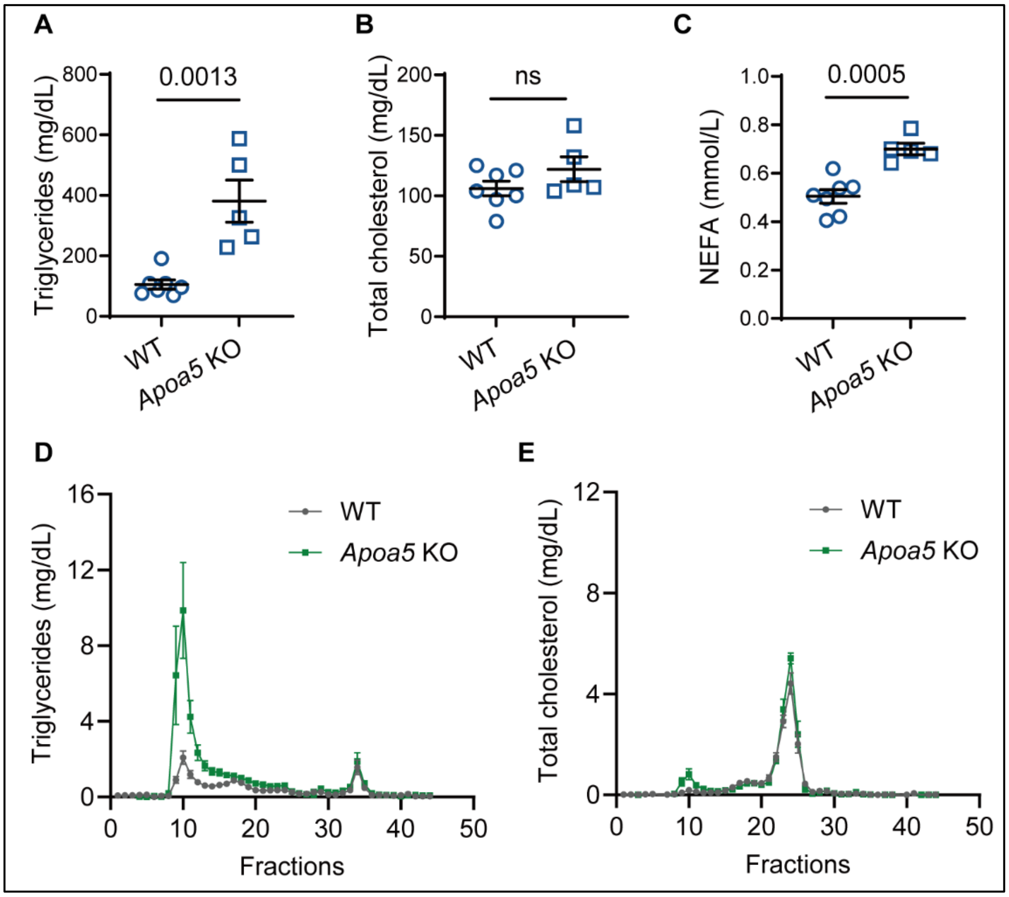


**Supplementary Fig. 2: Male *Apoa5*-deficient mice have higher plasma triglycerides and non-esterified fatty acids levels.**

10-12-week-old *Apoa5*-deficient mice and littermate controls were fed on a standard rodent laboratory diet and infused subcutaneously with AngII (1,000 ng/kg/min) for 2 weeks. The plasma was then collected. **A-C**, plasma triglycerides (**A**), cholesterol (**B**), and non-esterified fatty acids (**C**) levels from two groups. **D-E**, triglycerides (**D**) and cholesterol (**E**) concentrations of size exclusion chromatography fractionated plasma from animals (n = 3 per group). Mann-Whitney U test for **A**; Student's t-test for **B, C**.

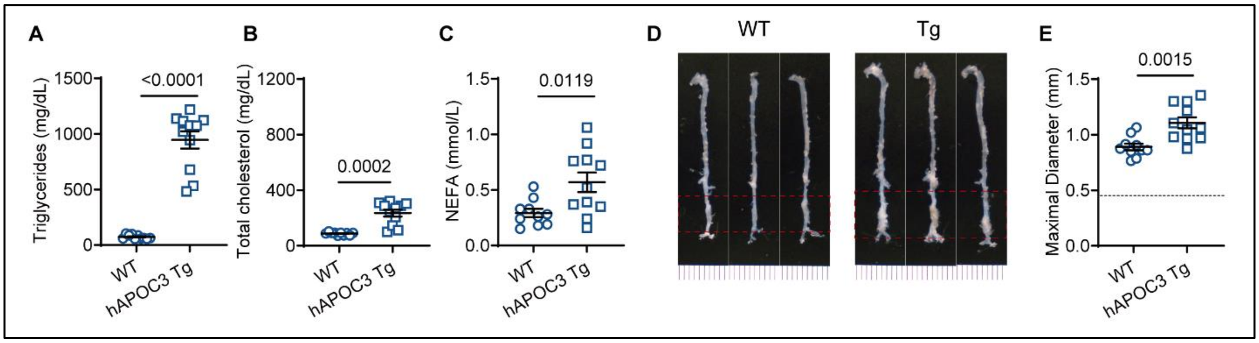


**Supplementary Fig. 3: Increased triglyceride concentrations accelerated AAA growth in male human *APOC3* transgenic mice in the porcine pancreatic elastase model.**

To induce AAA, porcine pancreatic elastase was applied to the infrarenal abdominal aorta of eight-week-old male h*APOC3* Tg mice or littermate WT controls. After 14 days, the AAA was evaluated, and blood samples were collected. **A**, Plasma triglycerides. **B**, Plasma total cholesterol. **C**, Plasma non-esterified fatty acids. **D**, Representative aortic tree. **E**, Maximal abdominal aortic diameter, dash line as the mean value of measured diameters from 7 sham-operated mice. Statistical analyses: Mann-Whitney U test for **A**; Student's t-test for **B**, **C**, **E**.

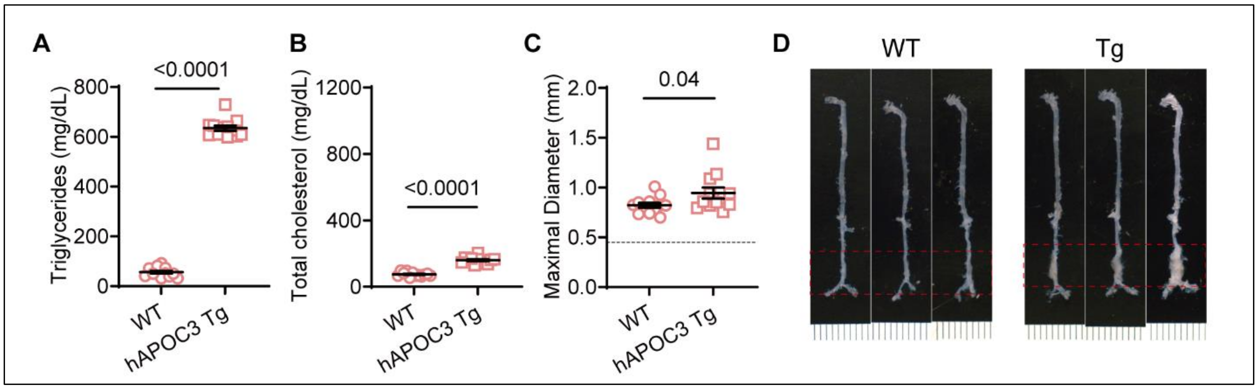


**Supplementary Fig. 4: Increased triglyceride concentrations accelerated AAA growth in female human *APOC3* transgenic mice in the porcine pancreatic elastase model.**

To induce AAA, porcine pancreatic elastase was applied to the infrarenal abdominal aorta of eight-week-old female h*APOC3* Tg mice or littermate WT controls. After 14 days, the AAA was evaluated and blood samples were collected. **A**, Plasma triglycerides. **B**, Plasma total cholesterol. **C**, Maximal abdominal aortic diameter, dash line is the mean value of measured diameters from 5 sham-operated mice. **D**, Representative aortic tree. Statistical analyses: Student's t-test for **A**, **B**. Mann-Whitney U test for **C**.

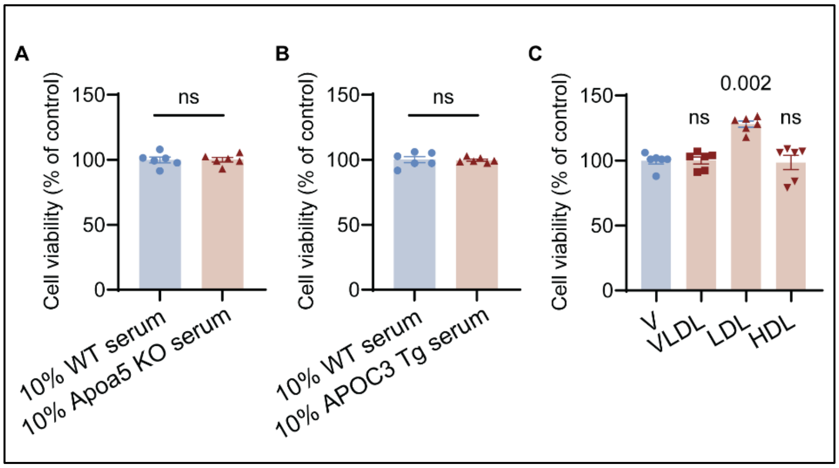


**Supplementary Fig. 5: Effects of serum from *Apoa5*-deficient mice, human *APOC3* transgenic mice, and lipoproteins on the cell viability of human aortic smooth muscle cells**

Human aortic smooth muscle cells (HASMCs) at a confluence of 80% were starved in OptiMEM-reduced serum medium for 24 hours, followed by treatment of 10% serum from *Apoa5* deficient mice or h*APOC3* transgenic mice for another 24 hours, 10% serum from littermates WT mice was used as control (**A**, **B**); or treat with 15 ug/ml VLDL, 50 ug/ml LDL, and 50 ug/ml HDL, with PBS as control (**C**). Student's t-test for **A**, **B**, One-way ANOVA followed by Sidak post hoc analysis for **C**.

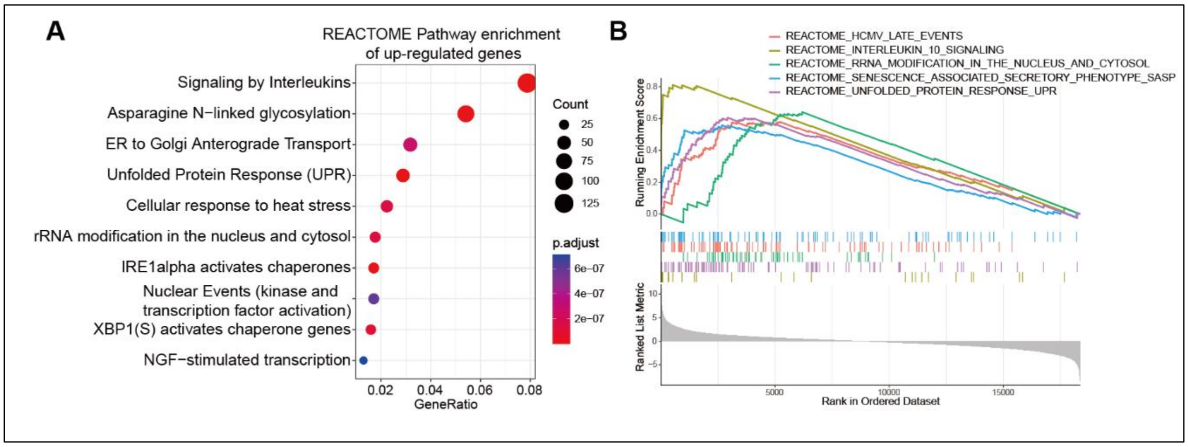


**Supplementary Fig. 6: Functional enrichment analysis of the upregulated genes in human aortic smooth muscle cells treated with 250 μM palmitic acid.**

**A**, Reactome pathway enrichment analysis. **B**, Gene Set Enrichment Analysis (GSEA), shown are the top five upregulated terms.

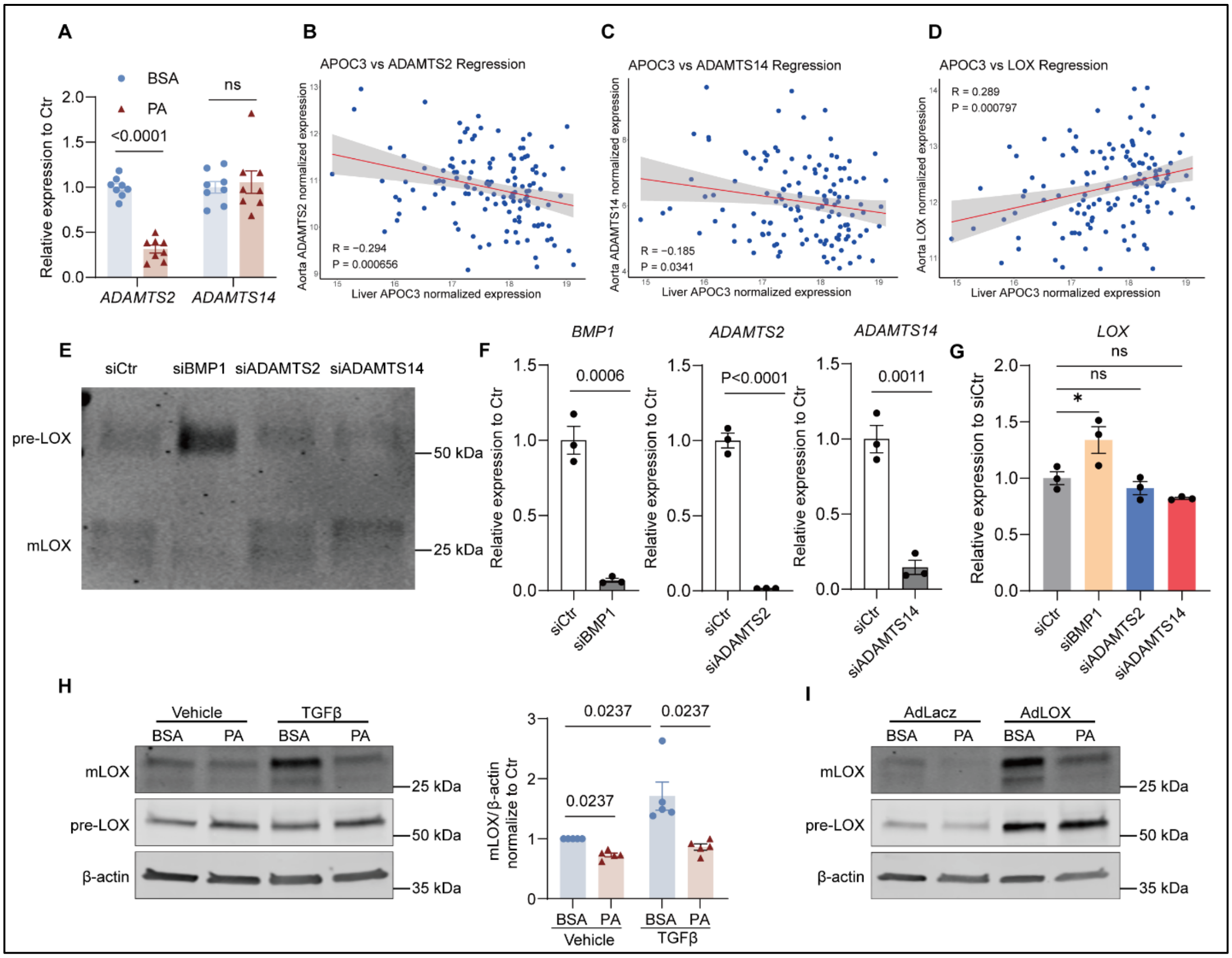


**Supplementary Fig. 7: Palmitic acid inhibits LOX maturation in HASMC**

**A**, RT-qPCR analysis of *ADAMTS2* and *ADAMTS14* in HASMC after 24-hours palmitic acid (PA) treatment. **B-D**, Correlation between liver *APOC3* expression level and aortic *ADAMTS2* (**B**), *ADAMTS14* (**C**), and *LOX* (**D**) expression level among 131 donors in the GTEx project. **E-G**, HASMC at 80% confluence were transfected with siBMP1, siADAMTS2, siADAMTS14, or scrambled siCtr at 16 nM for 48 hours, then refreshed with OptiMEM for another 24 hours. The conditioned medium was used for Western blot, and whole cells were used for RNA extraction and qPCR. **E**, Representative WB of LOX in the conditioned medium. **F**, qPCR shows effective knockdown efficiency. **G**, qPCR shows relative expression of *LOX*. **H**, HASMCs were starved in OptiMEM-reduced serum medium for 24 hours, then incubated with PA (250 μM) or vehicle along with TGF-β (10 ng/ml) or vehicle for another 24 hours. Total proteins from cell were extracted for Western blot analysis. Shown are representative Western blot images of the mature and premature forms of LOX in the cell lysates (left) and quantification analysis of mature LOX protein abundance (data from 5 independent experiments). **I**, HASMCs were transfected with adenovirus LacZ or LOX (30 MOI) for 2 hours in a growth medium, then starved in OptiMEM for another 22 hours. Cells were incubated with PA (250 μM) or vehicle for an additional 24 hours. Total proteins from cell were then extracted for Western blot analysis of mature and premature LOX. Student's t-test for **A**,

**F**; One-way ANOVA followed by Sidak post hoc analysis for **G;** Mann-Whitney U test followed by Bonferroni correction for **H**.

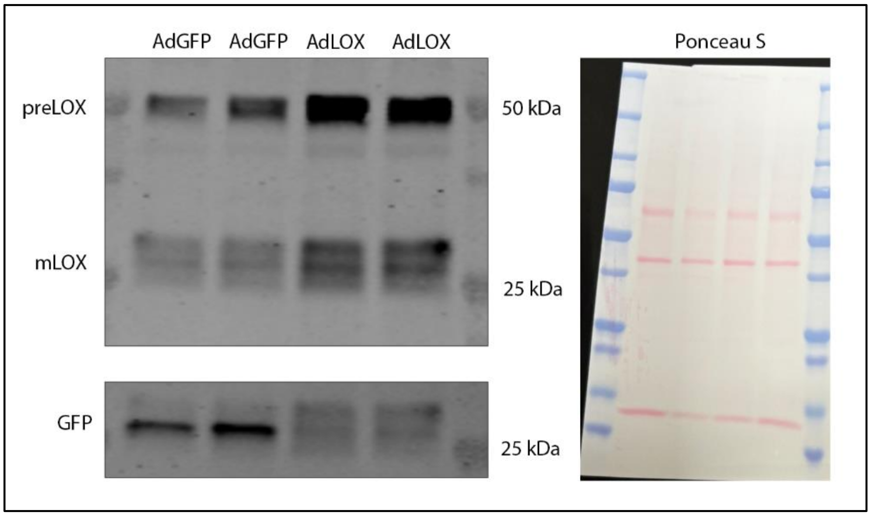


**Supplementary Fig. 8: Local adenovirus LOX overexpression increases mature LOX in the suprarenal abdominal aorta.**

$9 \times 10^8$ pfu of adenovirus expressing GFP or LOX were delivered to the suprarenal abdominal aorta in 10-week-old wide-type mice. 6 days later, the mice were euthanized, suprarenal abdominal aortas were isolated, and total protein was extracted and analyzed by Western blot to detect GFP and LOX abundance in suprarenal abdominal aortas.

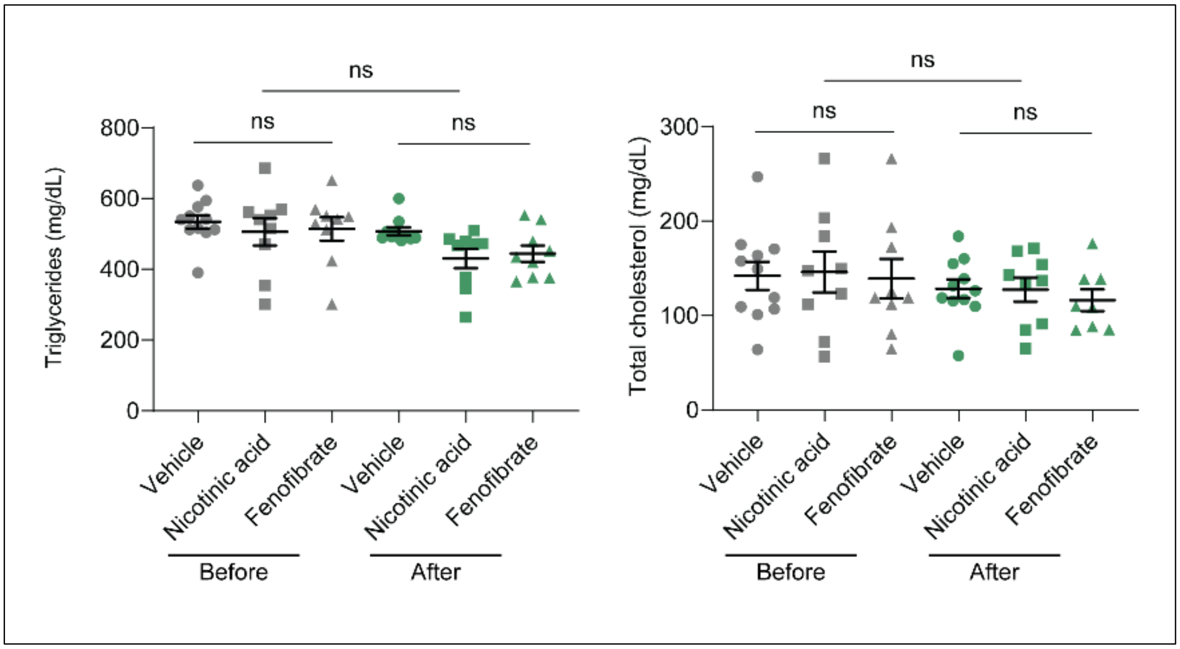


**Supplementary Fig. 9: Effects of nicotinic acid and fenofibrate on plasma triglycerides and total cholesterol in female *APOC3* transgenic mice.**

Eight-to-ten-week-old female *APOC3* transgenic mice were divided into three groups: fenofibrate at 60 mg/kg/day, Nicotinic acid at 1,000 mg/kg/day, and vehicle control (n = 9/group) by gavage. After two weeks of treatment, the plasma triglycerides (TG) and total cholesterol (TC) concentrations were measured. Data are presented as dots and Mean ± SEM. Kruskal-Wallis test followed by Dunn's post hoc analysis for TG and One-way ANOVA followed by Sidak post hoc analysis for TC.

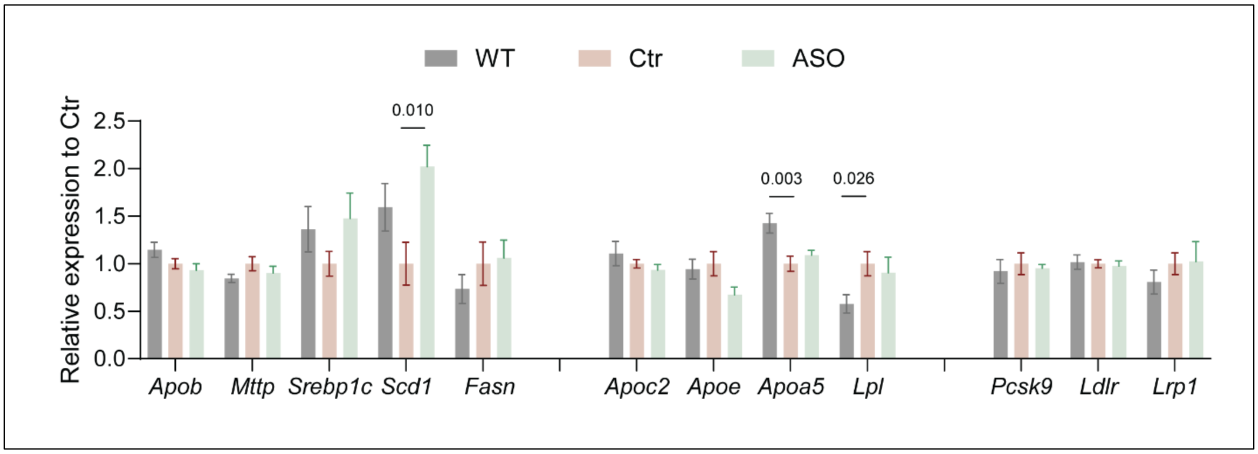


**Supplementary Fig. 10: mRNA expression among three groups in the *Angptl3* ASO study in the male h*APOC3* Tg mice.** One-way ANOVA followed by Sidak post hoc or Kruskal-Wallis test followed by Dunn's post hoc analysis for each gene. Only showing significant differences.

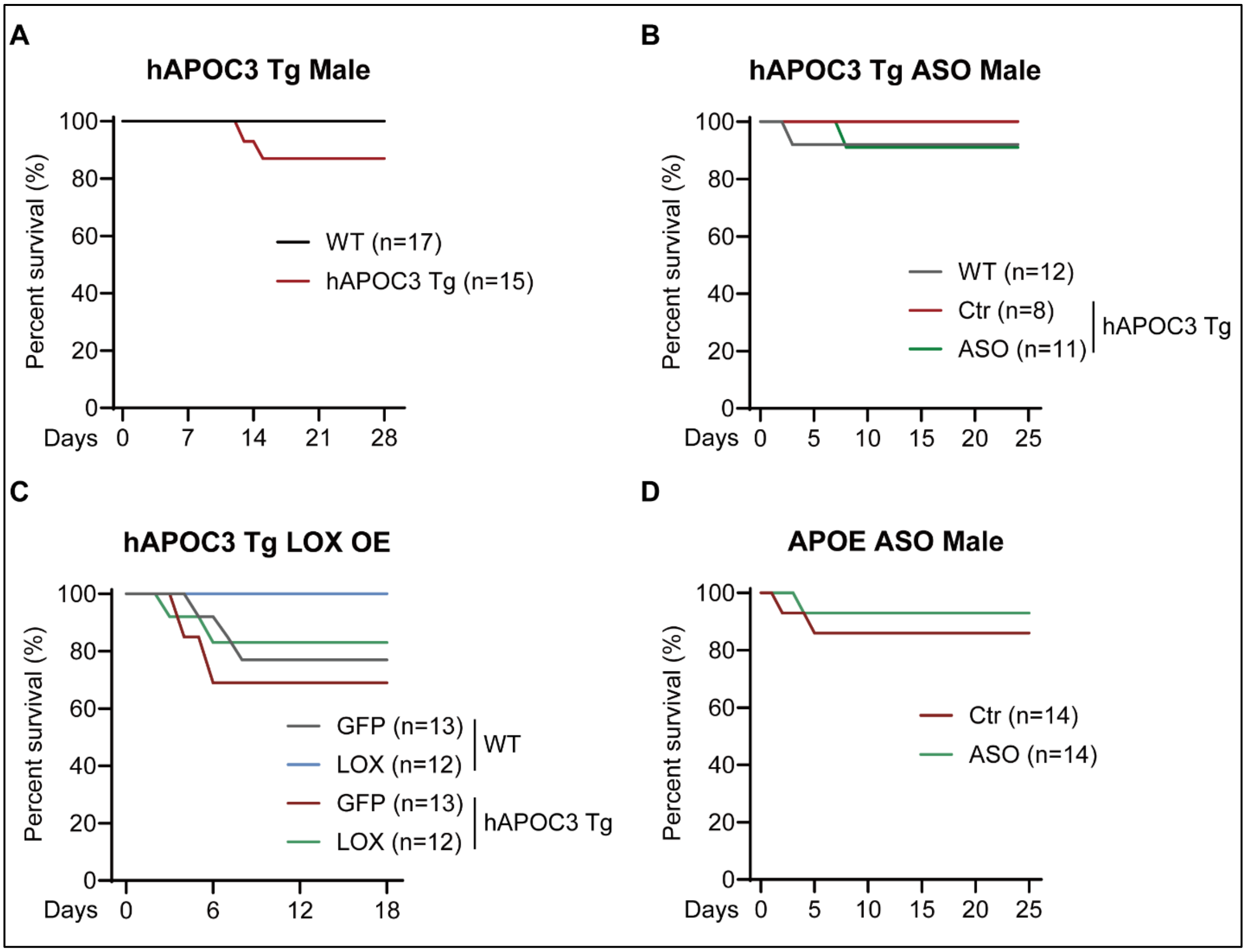


**Supplementary Fig. 11: Kaplan-Meier curve in the h*APOC3* Tg, h*APOC3* Tg ASO, h*APOC3* Tg LOX overexpression, and *Apoe*-deficient ASO animal cohorts.**

### Supplementary Tables

**Supplementary Table 1:** Full MR results of causal effects of 2,698 circulating proteins on the risk of AAA

**Supplementary Table 2:** Muti-variable MR results of causal effects of 41 circulating proteins on the risk of AAA

**Supplementary Table 3:** MR-BMA analysis to rank most likely causal exposures of AAA

**Supplementary Table 4:** Full MR results of causal effects of 233 circulating metabolites on the risk of AAA

**Supplementary Table 5:** Differentially expressed gene analysis of Palmitic acid vs vehicle treated HASMCs

**Supplementary Table 6:** Primers list for real-time PCR

**Supplementary Table 7:** Summary of plasma TG and TC in different cohorts
